# Pathways to suicidal ideation for young people engaged in mental health care

**DOI:** 10.1101/2025.02.11.25322101

**Authors:** Mathew Varidel, Ian B. Hickie, Victor An, Sally Cripps, Roman Marchant, Jo Robinson, Louise La Sala, William Capon, Ashlee Turner, Alexander Tashevski-Beckwith, Elizabeth Scott, Frank Iorfino

## Abstract

**Background:** Suicidal thoughts and behaviours (STBs) have a profound impact on individuals, communities, and healthcare systems. A wide range of factors have been shown to be associated with STBs. Within prior research it is also common to distinguish between proximal and distal factors, usually by distinction of short compared to long-term prediction. We frame the distinction between proximal and distal factors on suicidal ideation as direct or indirect dependencies using the inferred structure of probabilistic graphical models (PGMs).

**Methods:** We used cross-sectional data from a sample of 1020 help-seeking individuals aged 12-25 years from Australia that while engaged in a mental health care, contributed data to a digital platform. We inferred the posterior distribution of the dependency structure assuming both undirected PGMs and Bayesian networks (BNs). We then used the BN analysis to infer the causal effect that changing one variable has on another using a counterfactual query.

**Results:** We show that factors encompassing depressed mood, functional impairment, poor social connection, and psychosis-like experiences are proximal. Whereas experiencing a traumatic event, anxiety, insomnia, and unrefreshed sleep are distal factors. Proximal factors tended to have the greatest effect on suicidal ideation, while anxiety symptoms and experiencing a traumatic event were the most influential distal factors.

**Conclusions:** These relative timings of events and their effects on suicidal ideation could be used to understand the future likelihood of suicidal ideation, and aid planning of targeted interventions.

## Introduction

Suicidal thoughts and behaviours (STBs) have a profound impact on individuals, communities and healthcare systems. Suicide is the fourth leading cause of death globally for individuals aged 15-29 years (World Health Organization, 2021), and the leading cause of death for individuals aged 15-24 years in Australia (Australian Institute of Health and Welfare, 2024). Suicidal thoughts are problematic in and of themselves and are prevalent (Jobes & Joiner, 2019), with a 12-month prior prevalence rate of approximately 15% for adolescents worldwide, with variation between 8-21% depending on location and study (Biswas et al., 2020; Uddin et al., 2019). A recent meta-analysis suggested that lifetime prevalence rates for suicidal ideation was 19% for youths in Australia and New Zealand (Van Meter et al., 2023).

Due to the impact of STBs, significant resources have focused on understanding the associated risk factors (Knipe et al., 2022; Nock et al., 2008). These factors are varied and include prior STBs or self-harm (Iorfino et al., 2018; Moran et al., 2024), interpersonal functioning (Ammerman & Jacobucci, 2023; Chu et al., 2017; Franklin et al., 2017; Heapy et al., 2024), personal functioning (Iorfino et al., 2023a; Skinner et al., 2023), clinical diagnoses (Nock et al., 2008; Y. E. Xu et al., 2023), specific psychological symptoms and states (O’connor & Nock, 2014), sleep problems (Kearns et al., 2020), and sociodemographic factors. Most of these risk factors have been found for adult samples, although similar risk factors are often found for youth (Bakken et al., 2025; Bilsen, 2018). Specific risk factors for youth include parental relationships (Biswas et al., 2020; Consoli et al., 2013), their parents’ prior history of STBs and other clinical factors (Christiansen et al., 2011), and school connectedness (Bakken et al., 2025).

Many of these risk factors have been shown to be associated with suicidal ideation specifically, rather than suicide attempts (Klonsky et al., 2016; May & Klonsky, 2016). While suicidal ideation is a prerequisite of suicide attempts, only one third of people with suicidal ideation make a suicide attempt within their lifetime (Nock et al., 2008). Similar rates of suicidal ideation to attempts are often reported in youths (Han et al., 2009; Nock et al., 2013; Orri et al., 2020). Although, Uddin et al. (2019) found 12-month prevalence for suicidal ideation and attempts were comparable in a worldwide youth sample.

There are also several theories that aim to explain the causal pathways to STBs. These theories encompass the progression to suicidal ideation to attempts, while highlighting the distinction between risk factors for suicidal ideation and suicidal behaviours. For example, the interpersonal theory of suicide (IPTS) suggests that thwarted belongingness and burdensomeness cause hopelessness which in turn can lead to suicidal thoughts (Van Orden, Witte, Cukrowicz, Braithwaite, Selby, Joiner, et al., 2010). Suicidal thoughts plus acquired psychological capability and means then lead to suicidal behaviours. The three-step theory of suicide suggests that the primary precursor to suicidal ideation is psychological pain; where low levels of suicidal ideation require a level of hopelessness, strong ideation requires connectedness to be overwhelmed by their pain, and suicide attempts require both practical means and a psychological tolerance for suicide (Klonsky & May, 2015), similar to those of IPTS. The integrated motivation-volitional model of suicidal behaviour includes similar progressional steps from suicidal ideation to attempts, but encompasses lower-level risk factors (e.g., thwarted belongingness, burdensomeness) into higher-level psychological theories, including the diathesis-stress model, theories of planned behaviour, and the differential activation hypothesis (O’Connor & Kirtley, 2018). Further theoretical advancements have developed to understand the dynamics of STBs (Bryan et al., 2020; Rugo-Cook et al., 2021). All of these theories highlight the complex paths that can lead to STBs.

This wide array of associated factors raises the question around the relative timing and causal pathways to STBs. Hence, these factors are often split into proximal (i.e., recent predictors of STBs, e.g., recent mood) or distal factors (i.e., long-term predictors that may be causally further upstream of proximal factors, e.g., sociodemographic factors) (Klonsky et al., 2016; Knipe et al., 2022). Understanding these causal pathways would help to identify appropriate intervention and preventative targets, as well as their relative timings.

Probabilistic graphical models (PGMs) are an approach to gain insight into the dependency structure between variables within one’s data (Koller & Friedman, 2009; Maathuis et al., 2018). In PGMs, variables are represented as nodes and conditionally dependent variables are indicated by lines connecting nodes (i.e., edges). Undirected PGMs are a specific case of a PGM that make no claim about the directionality of edges between nodes, which have become commonplace in psychological research (Borsboom et al., 2021). They have been used to show the dependency structure of various factors with suicidal ideation, including theoretically considered factors (e.g., burdensomeness, thwarted belongingness; De Beurs et al., 2019), along with a range of psychological and behavioural factors (Holman & Williams, 2020; Jeon et al., 2024; Li & Kwok, 2023; Núñez et al., 2018).

Bayesian networks (BNs) are an alternative PGM to describe the dependency structure (Koller & Friedman, 2009). BNs represent the dependency structure with a directed acyclic graph (DAG), that has directed edges that if followed cannot result in a cycle. A potential advantage of BNs over undirected PGMs is that they allow for causal interpretations (Dablander & Hinne, 2019; Ryan et al., 2022), thus providing a data-driven approach to causal hypothesis generation and causal effect estimation. BNs can be learnt from observed data up to an equivalence class even from cross-sectional data. A recent paper used BNs to understand the dependencies between psychosocial factors and suicidal ideation, where they found that suicidal ideation was dependent on total depressive symptoms, total anxiety symptoms, self-efficacy, resilience, and quality of life (Delgadillo et al., 2023). In this paper, we use undirected PGMs and BNs to further our understanding of the potential pathways to STBs using a range of clinical and psychosocial variables for young people that present to services across Australia.

## Methods

This study was approved by the Northern Sydney Local Health District Human Research Ethics Committee (HREC/17/HAWKE/480) and all participants gave online informed consent (via an opt-out process).

### Participants

Participants were a group of young people aged 12-25 years who presented to *headspace* services in Australia (McGorry et al., 2007) between November 2018 and October 2023 and used the Innowell platform (Iorfino et al., 2019). Individual’s must have completed all item-level questions that appear within this paper to have been admitted to the study.

### Innnowell

Innowell is a digital technology used clinically for the assessment, management, and monitoring of mental health and well-being. The web-based platform allows individuals to complete a multidimensional clinical assessment at entry into care and over the course of care in a self-directed or clinically directed way. Results are displayed on a personalised dashboard to provide a better understanding of an individual’s needs, track progress, and get access to recommended self-directed or clinical care options. The tool is not intended as a crisis management tool, although a standardised notification is used for those young people reporting suicidal thoughts and behaviours, so that an immediate clinical response and protocol can be engaged by the service.

### Measures

The initial questionnaire in the Innowell platform covers multiple domains. The domains that we use in this analysis are; 1) mental health, including; psychosis-like experiences using the 16-item Prodromal Questionnaire (PQ16; Ising et al., 2012); depressed mood using the Quick Inventory of Depression Symptomatology (QIDS; Rush et al., 2003); anxiety using the Overall Anxiety Severity and Impairment Scale (OASIS; Norman et al., 2006); mania-like experiences using the Altman Self-Rating Mania Scale (ASRM; Altman et al., 1997), 2) functioning impairment due to mental health using the Work and Social Adjustment Scale (WSAS; Mundt et al., 2002), 3) social connection using Schuster’s Social Support Scale (SSSS; Schuster et al., 1990), 4) lifetime experience of a traumatic event, 5) sleep quality and timings using the BMC Sleep-Wake Cycle (Buysse et al., 1989; Roenneberg et al., 2003; Vernon et al., 2010; Zhang et al., 2013), and 6) suicidal ideation using the Suicidal Ideation Attributes Scale (SIDAS; Van Spijker et al., 2014). We also account for demographic variables including age, sex, and whether they presented to an urban or regional service (Capon et al., 2023). These questions probed those domains over approximately the last four weeks. We also used a question regarding suicide attempts in the past three months from the Columbia-Suicide Severity Rating Scale (C-SSRS; Posner et al., 2011). See Supplementary Materials 1 for further details.

## Statistical analyses

All statistical analyses were performed in R version 4.4.1 (R Core Team, 2024).

### Differential Odds of Suicidal Thoughts and Behaviours

We estimated the relative change in odds of having an STB outcome conditional on a change in a sample characteristic. Two different binary outcomes are considered; 1) those with or without a recent suicide attempt, or 2) those with or without high suicidal ideation (SIDAS ³ 20). This is estimated using Bayesian logistic regression implemented in the rstanarm package (Goodrich et al., 2024). Marginal odds-ratios were estimated only including the clinical, psychosocial, and demographic factor as a predictor of the STB outcome. Conditional odds-ratios were estimated by including all factors as predictors. We report the odds ratio posterior median and 95% highest density credible interval (Makowski et al., 2019).

### Undirected Probabilistic Grapical Model

We estimated the posterior distribution for a Gaussian graphical model (GGM). A GGM is an undirected PGM that assumes the data follow a multivariate normal distribution. This inference was performed using BDgraph (Mohammadi & Wit, 2019). BDgraph uses a birth-death Markov chain Monte Carlo scheme with proposals that add and remove edges. We ran four chains for five million iterations each with the first thousand iterations removed as burnin. As a convergence check, we ensured that the difference in the estimated edge probabilities across all chain pairs was within 10%. Note that for all inferences for PGMs we removed items that were deemed to overlap with other questionnaires.

### Bayesian Network

Inference for the posterior distribution of Bayesian Networks was then performed. Estimating the posterior distribution of BNs is difficult due to the large number of possible DAGs. For example, the number of possible DAGs for a 10-node BN is on the order of 10^18^. Several simplifications were implemented to reduce the possible number of DAGs. We collapsed the items into factors in accordance with prior research. PQ16 was collapsed into unusual thoughts and perceptional abnormalities (Ising et al., 2012). SSSS was collapsed into a perceived negative and a perceived lack of positive social connection with family and friends (Schuster et al., 1990). QIDS was collapsed into insomnia (sum of questions 1-3), hypersomnia (question 4), motor activation (sum of questions 15 and 16), recent appetite and weight change (max of QIDS_WEIGHT and QIDS_APPETITE) with the remaining questions separated. This broadly follows the original aggregated items by Rush et al. (2003), with the exception that we split out insomnia and hypersomnia and tended to sum rather than calculate the max, so that we could retain information from across all items. OASIS, WSAS, and SIDAS were considered as one factor for each questionnaire representing anxiety, functioning, and suicidal ideation respectively, with the total scores used to estimate the BN. Further detail can be found in Supplementary Table S1.

We also made the following assumptions with respect to the pairwise edges. Suicidal ideation could not be a parent of any other nodes, thus assuming that it is an outcome. A traumatic event could only be a child node of age and sex. Unrefreshed sleep could not be a parent of insomnia or hypersomnia nodes. Also, as WSAS asks about functioning conditional on an individual’s mental health it also could not be a parent of mental health factors. We also assume that age and sex could not be children of any other node.

Posterior sampling was achieved using a bespoke implementation of the Partition Markov chain Monte Carlo (PMCMC) scheme (Kuipers & Moffa, 2017; Varidel, 2024). We started all chains from an optimised starting point as estimated by taking the *maximum a posterior* estimate over 10^4^ runs of tabu search, where each run was started from random locations (Scutari, 2010; Scutari et al., 2019). We ran eight chains of PMCMC for 10^7^ iterations saving every 500^th^ iteration, leading to 20000 samples. The split-*R̂* convergence statistic was *R̂* < 1.01 for the log partition score with *N*_eff_ = 6645 effective samples (Gelman et al., 2013; Vehtari et al., 2021). For the DAG scores, we found *R̂* < 1.01 with *N*_eff_ = 1081. We also checked the concordance of the mean pairwise edge probabilities (Suter et al., 2023), where the difference between all edge probabilities between all chains was always < 20%. These statistics suggest reasonable convergence and resolution of the chains.

### Causal effects

We estimated the average causal effects (ACE) across our sample that intervening to deterministically change a node would have on another node. The ACE can be thought of as the typical difference of the effect node if we changed the intervened node deterministically. Thus, ACE is a counterfactual query of the Bayesian network, where the posterior distribution for the ACE of *A* on *B* is given by,

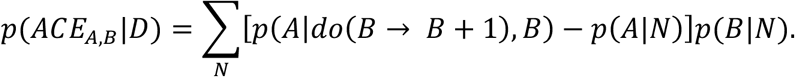

This quantity is marginalised over the posterior distribution of the BN *N* given the data *D*. As *p*(*A*|*do*(*B* → *B* + 1), *N*) is a causal estimand due to an hypothesised intervention, we must correct for the confounders of *A* and *B* (Pearl et al., 2016), which is implemented in the Bestie package (Kuipers & Moffa, 2022). Marginalisation over the BNs is implemented by performing this calculation per iteration from our posterior sample of BNs. The ACE posterior distribution is typically multimodal as when there is no path from intervened node *A* to the effect node *B* the effect size is zero, and non-zero when there is a path. As such, we chose to report the median and the 25% and 75% quantiles. When zero is not within the interquartile range the probability of a path from *A* to *B* will be at least 50%.

## Results

### Differential Odds of Suicidal Thoughts and Behaviours

The sample comprised of 1020 individuals (72.0% female, 81.8% urban) with a mean age of 19.6 years (SD, 2.8 years). Of these individuals, 21 (2.1%) had a suicide attempt within the last three months. The marginal odds of having a recent suicide attempt increased with experiencing a traumatic event (OR, 2.80, 95% HDI, 1.08, 7.96), female sex (OR, 4.01, 95% HDI, 1.12, 21.43), depressed mood (OR, 1.35, 95% HDI, 1.19, 1.52), suicidal ideation (OR, 1.21, 95% HDI, 1.15, 1.28), psychosis-like experiences (OR, 1.21 , 95% HDI, 1.09, 1.35), anxiety (OR, 1.16, 95% HDI, 1.04, 1.32), functioning (OR, 1.07, 95%, 1.01, 1.13), and potentially social connection (OR, 1.12, 95% HDI, 1.00, 1.28). However, the conditional odds-ratios only increased with suicidal ideation (OR, 1.26, 95% HDI, 1.18, 1.36) and potentially lowered by personal function (OR, 0.90, 95% HDI, 0.82, 0.99). This result suggests that suicidal ideation is the only proximal factor on a recent suicidal attempt. Further details are shown in Table 1 with more detail provided in Supplementary Tables S3-S5.

**Table 1.**
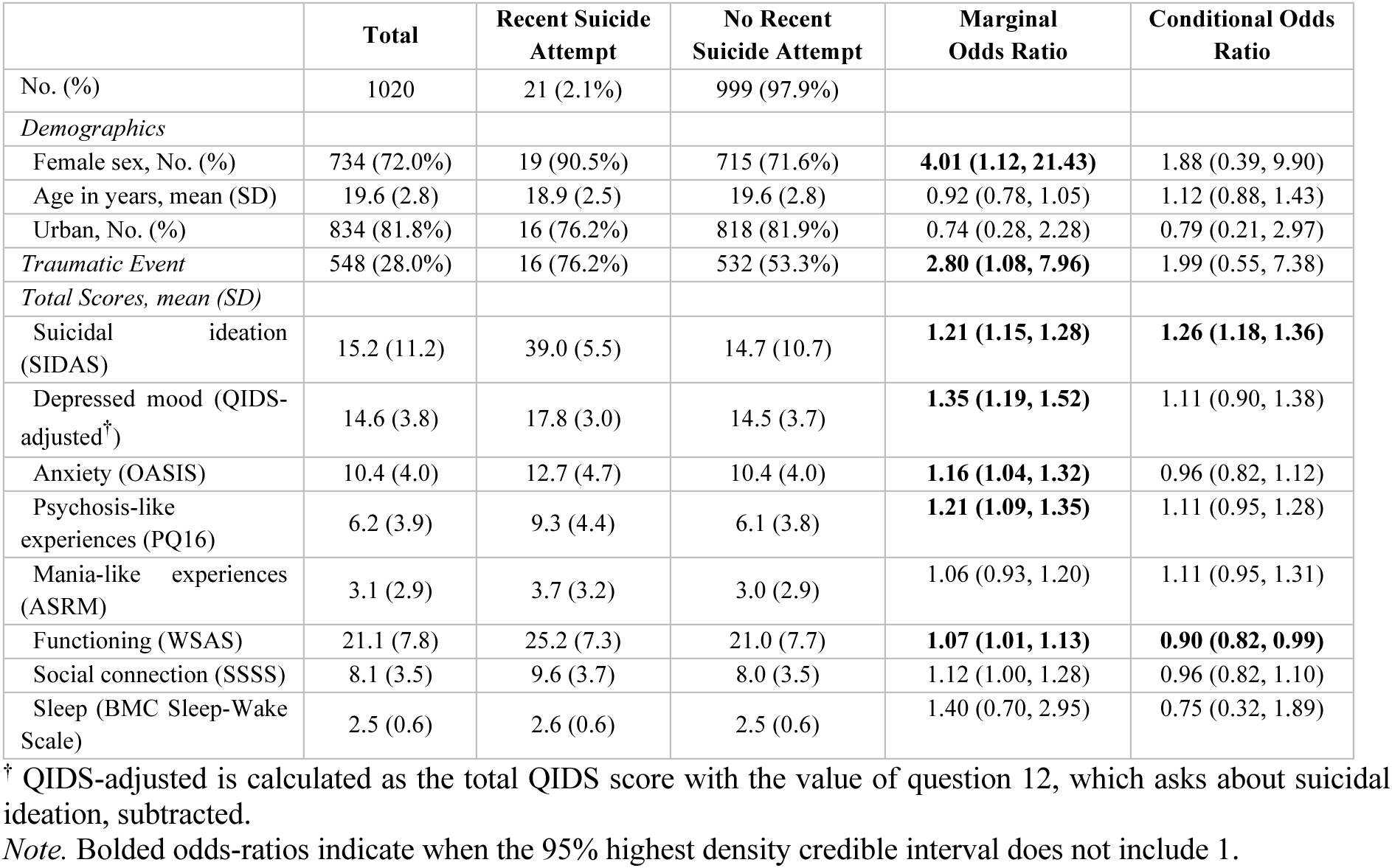
Sample characteristics and odds-ratios for a recent suicide attempt. Marginal odds-ratios estimate the unconditioned association for a recent suicide attempt given the sample characteristic. Conditional odds-ratio estimate the association between the sample characteristic and the outcome while controlling for all other sample characteristics. We report the median and 95% highest density interval for the sample characteristic.

There were 321 (31.5%) individuals within the high suicidal ideation category (SIDAS ≥ 20). The marginal odds of having high suicidal ideation increased with experiencing a traumatic event (OR, 1.49, 95% HDI, 1.14, 1.97), depressed mood (OR, 1.25, 95% HDI, 1.20, 1.31), sleep (OR, 1.43, 95% HDI, 1.14, 1.76), social connection (OR, 1.14, 95% HDI, 1.10, 1.19), anxiety (OR, 1.14, 95% HDI, 1.10, 1.18), psychosis-like experiences (OR, 1.14, 95% HDI, 1.10, 1.18), and functioning (OR, 1.11, 95% HDI, 1.09, 1.14). Whereas being older tended to decrease the odds of having high suicidal ideation (OR, 0.93, 95% HDI, 0.89, 0.98). Conditional odds ratios suggested that the proximal characteristics that increased high suicidal ideation were depressed mood (OR, 1.13, 95% HDI, 1.07, 1.21), psychosis-like experiences (OR, 1.06, 95% HDI, 1.01, 1.11), functioning (OR, 1.08, 95% HDI, 1.05, 1.11) and potentially social connection (OR, 1.05, 95% HDI, 1.00, 1.00), with older age still reducing the odds (OR, 0.91, 95% HDI, 0.86, 0.97). Further details are shown in Table 2 with further information provided in Supplementary Tables S6-S8. We found no association between mania-like experiences and being in an urban location with either a recent suicide attempt or being in the high suicidal ideation category, thus we do not consider these questions within subsequent analyses.

**Table 2.** Sample characteristics and odds-ratios for high suicidal ideation (SIDAS ≥ 20). Marginal odds-ratios estimate the unconditioned odds-ratio for high suicidal ideation given the sample characteristic. Conditional odds-ratio estimate the association between the sample characteristic and the outcome while controlling for all other sample characteristics. We report the median and 95% highest density interval for the sample characteristic.

|  | Total | High Suicidal Ideation | Low Suicidal Ideation | Marginal Odds Ratio | Conditional Odds Ratio |
| --- | --- | --- | --- | --- | --- |
| No. (%) | 1020 | 321 (31.5%) | 699 (68.5%) |  |  |
| *Demographics* |  |  |  |  |  |
| Female sex at birth, No. (%) | 734 (72.0%) | 242 (75.4%) | 492 (70.4%) | 1.29 (0.96, 1.78) | 1.07 (0.77, 1.52) |
| Age in years, mean (SD) | 19.6 (2.8) | 19.2 (2.7) | 19.7 (2.8) | **0.93 (0.89, 0.98)** | 0.91 (0.86, 0.97) |
| Urban, No. (%) | 834 (81.8%) | 254 (79.1%) | 580 (83.0%) | 0.78 (0.55, 1.09) | 0.95 (0.64, 1.40) |
| Traumatic Event | 548 (53.7%) | 194 (60.4%) | 354 (50.6%) | **1.49 (1.14, 1.97)** | 0.97 (0.71, 1.33) |
| *Total Scores, mean (SD)* |  |  |  |  |  |
| Depressed mood (QIDS-adjusted<sup>†</sup>) | 14.6 (3.8) | 16.5 (3.4) | 13.7 (3.6) | **1.25 (1.20, 1.31)** | **1.13 (1.07, 1.21)** |
| Anxiety (OASIS) | 10.4 (4.0) | 11.7 (4.1) | 9.8 (3.8) | **1.14 (1.10, 1.18)** | 0.98 (0.93, 1.02) |
| Psychosis-like experiences (PQ16) | 6.2 (3.9) | 7.5 (4.0) | 5.6 (3.7) | **1.14 (1.10, 1.18)** | **1.06 (1.01, 1.11)** |
| Mania-like experiences (ASRM) | 3.1 (2.9) | 3.0 (3.0) | 3.1 (2.8) | 0.98 (0.94, 1.03) | 0.96 (0.91, 1.01) |
| Functioning (WSAS) | 21.1 (7.8) | 25.0 (7.1) | 19.3 (7.4) | **1.11 (1.09, 1.14)** | **1.08 (1.05, 1.11)** |
| Social connection (SSSS) | 8.1 (3.5) | 9.2 (3.7) | 7.5 (3.3) | **1.14 (1.10, 1.19)** | 1.05 (1.00, 1.10) |
| Sleep (BMC Sleep-Wake Scale) | 2.5 (0.6) | 2.6 (0.6) | 2.5 (0.7) | **1.43 (1.14, 1.76)** | 0.93 (0.72, 1.19) |

<sup>†</sup> QIDS-adjusted is calculated as the total QIDS score with the value of question 12, which asks about suicidal ideation, subtracted.
*Note.* Bolded odds-ratios indicate when the 95% highest density credible interval does not include 1.

### Undirected probabilistic graphical model

The inferred item-level undirected PGM is dense with suicidal ideation being directly or indirectly associated with all domains (Figure 1). As would be expected, we see dependencies between items within each question set. Items in depressed mood, functioning, and anxiety are also highly dependent. Suicidal ideation is directly associated with all domains except anxiety and experiencing a traumatic event (PTSD5_EVENT). Nodes that bridge the relationship between their respective domain and suicidal ideation are sadness (QIDS_5), self-perception (QIDS_11), concentration (QIDS_10), uncaring (SSSS_1) and unavailable (SSSS_6) friends and family, hearing voices (PQ16_13), a lack of control of thoughts (PQ16_11), and home management (WSAS_2). See further quantitative detail in Supplementary Figures S1-S3.

**Figure 1.**
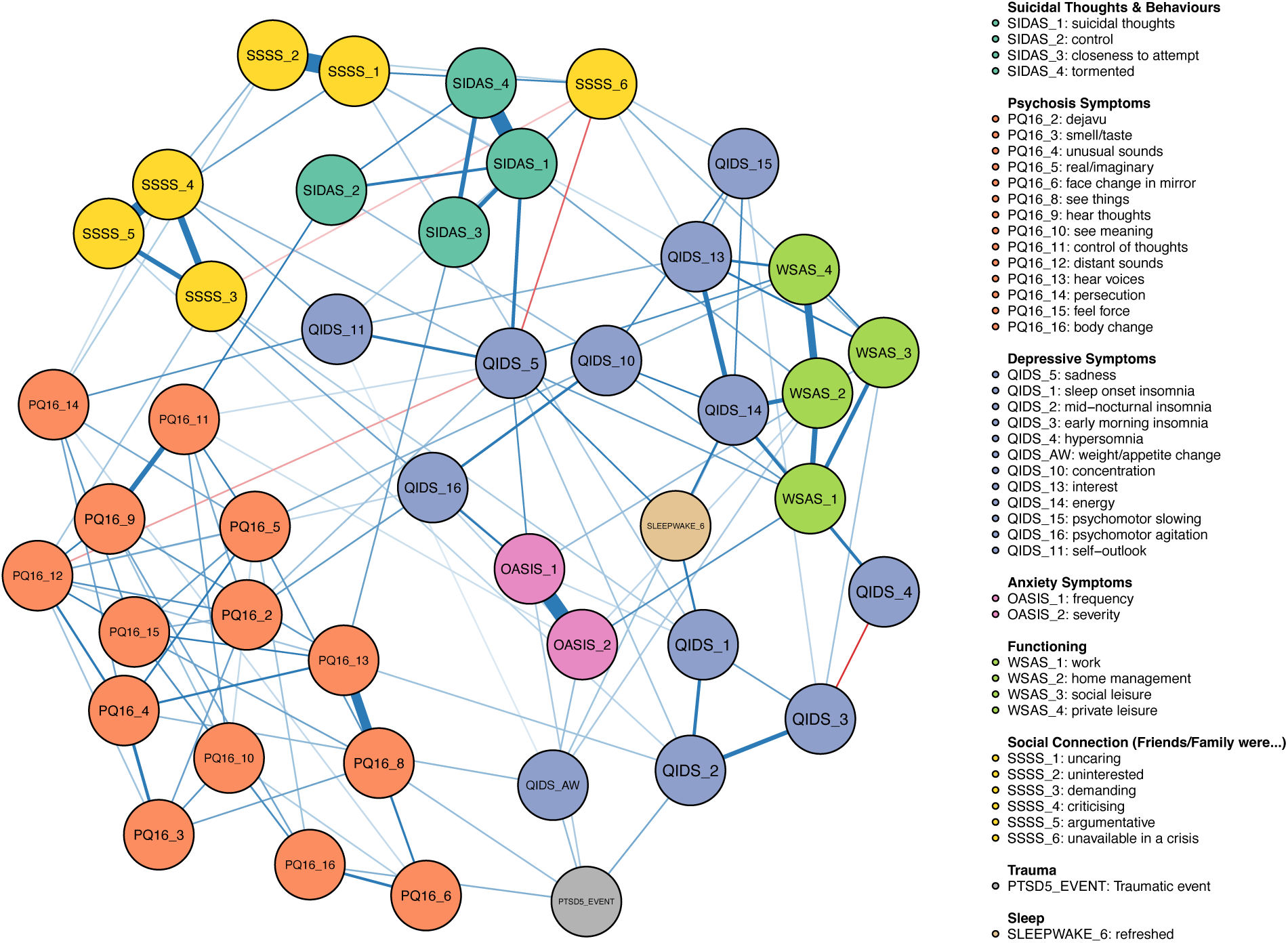
Item-level undirected probabilistic graphical model. Edges are shown with *p* ≥ 0.5.

### Bayesian network

We show the *maximum a posteriori* estimate for the subgraph that includes suicidal ideation along with its ancestors in Figure 2. This result shares similarities with the inferred undirected PGM. Suicidal ideation was directly dependent on sadness, functioning, lack of positive social connection, and unusual thoughts. For simplicity, we will refer to nodes that suicidal ideation is directly dependent on as ‘proximal factors’. Upstream of the proximal factors we see that suicidal ideation is indirectly dependent on several factors. We will refer to the nodes that have indirect paths to suicidal ideation as ‘distal factors’. However, these distinctions should be considered relative rather than absolute. See further quantitative detail in Supplementary Figures S4-S7.

**Figure 2.**
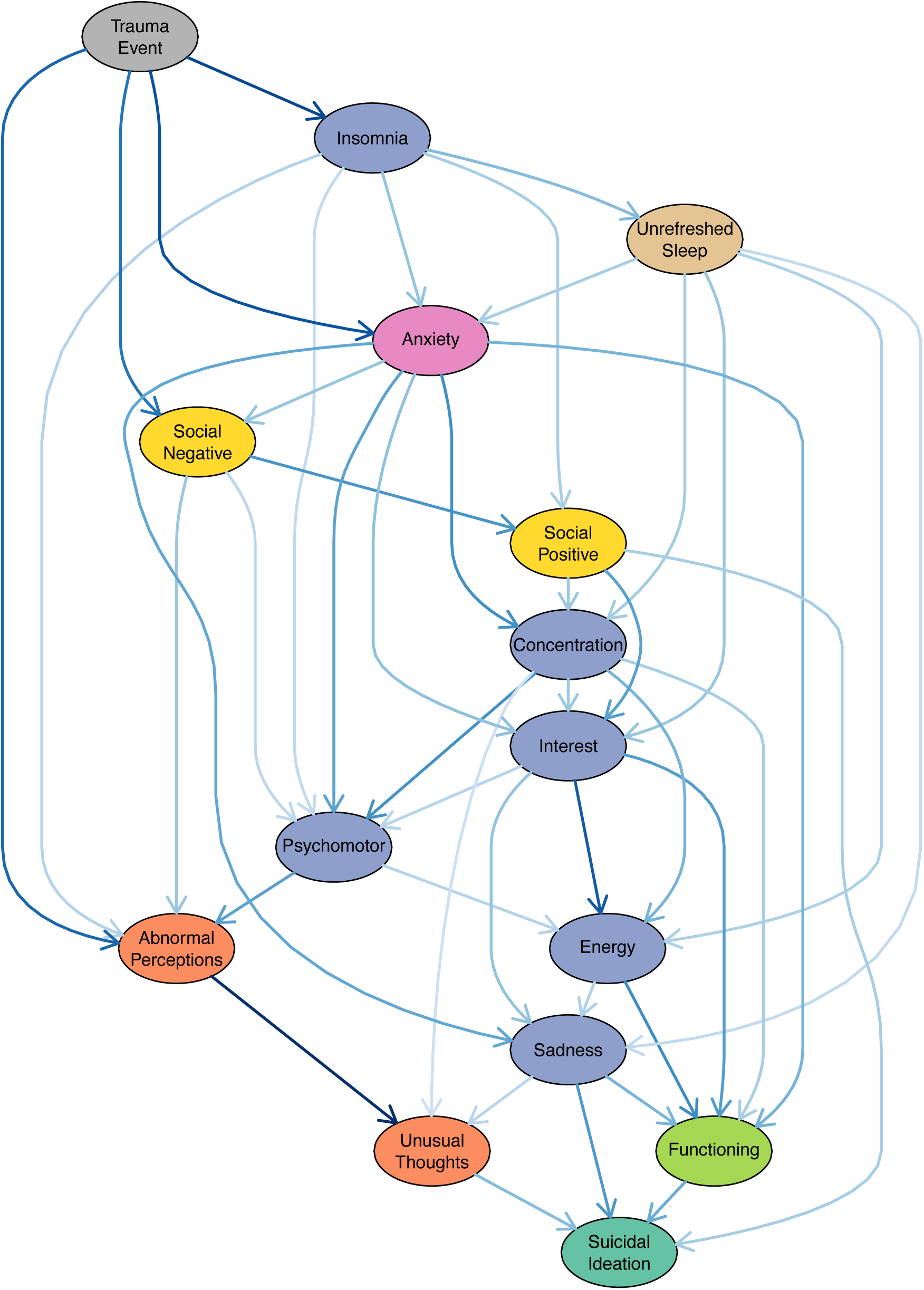
Factor-level *maximum a posteriori* directed acyclic subgraph, where we only show suicidal ideation and its ancestor nodes. All pairwise edges within this subgraph have posterior probability > 0.5. Edges are coloured in darker blue in accordance with the estimated value for the linear regression coefficients given the subgraph.

### Causal effects

All question sets have factors with positive average causal effects (ACE) on suicidal ideation. The greatest ACEs on suicidal ideation are from sadness (ACE, 0.30, IQR, 0.25, 0.36), functioning (ACE, 0.20, IQR, 0.14, 0.26), unusual thoughts (ACE, 0.18, IQR, 0.12, 0.24), anxiety (ACE, 0.16, IQR, 0.11, 0.20), perceived lack of positive interactions with family and friends (ACE, 0.16, IQR, 0.09, 0.23), experiencing a traumatic event (ACE, 0.15, IQR, 0.09, 0.19), interest (ACE, 0.14, IQR, 0.07, 0.19), and abnormal perceptions (ACE, 0.12, IQR, 0.08, 0.17).

Nodes appearing early in the topological ordering including experiencing a traumatic event, insomnia, unrefreshed sleep, and anxiety symptoms have wide ranging effects. For example, experiencing a traumatic event has strong effects on many factors including abnormal perceptions (ACE, 0.50, 0.39, 0.62), anxiety (ACE, 0.39, IQR, 0.27, 0.52), unusual thoughts (ACE, 0.37, IQR, 0.29, 0.49), insomnia (ACE, 0.36, IQR, 0.21, 0.48), and perceived negative interactions with family and friends (ACE, 0.36, IQR, 0.13, 0.49). See Figure 3 for the median values for cases where zero is not within the IQR. Further detail including the IQR along with sampling statistics for those edges with IQR inconsistent with zero showing reasonable convergence (maxF*R̂*G = 1.01) and resolution (min(*N*_eff_) = 3219) can be found in Supplementary Table S8.

**Figure 3.**
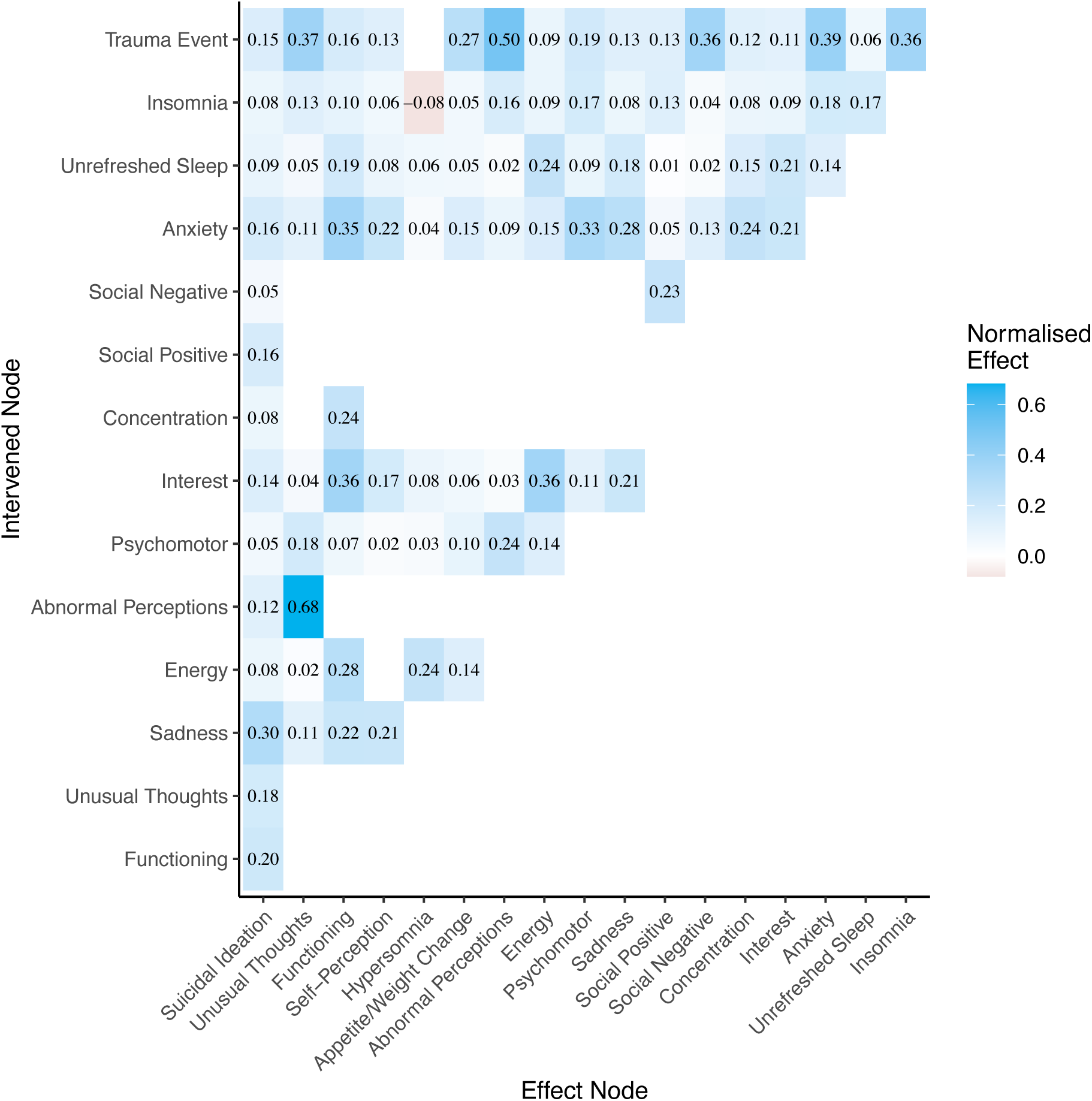
Factor-level average causal effects. This shows the median change in the effect nodes on a normalised scale given a 1-sigma change for all variables, except for experiencing a traumatic event which corresponds to a category change. We only show causal effects where zero is not within the IQR.

## Discussion

This work evaluated the dependencies for a range of clinical and psychosocial factors on STBs for those aged 12-25 years in Australia that while engaged in a mental health care, contributed data to a digital platform. Our results consistently revealed that depressive symptoms (particularly sadness), functional impairment, poor social connection (particularly a perceived lack of positive support), and psychosis-like experiences (particularly through hearing voices and a lack of control of thoughts in the unusual thoughts factor) were proximal factors to suicidal ideation, with most other clinical and psychosocial factors being distal. This wide array of influencing factors on suicidal ideation provides evidence for comprehensive youth care models (Hickie et al., 2019; McGorry et al., 2024), that include the consideration for a wide array of clinical, psychosocial, and comorbidity factors.

Distinguishing proximal and distal factors on suicidal ideation allows for a better understanding of the relevant information required to understand, predict, and prevent STBs. Proximal factors provide the most information about risk of suicidal ideation, as suicidal ideation is conditionally independent of distal factors given knowledge about proximal factors. Thus, given knowledge about an individual’s proximal factors, there is little to no extra predictive information gained by understanding the distal factors. On the other hand, understanding an individual’s distal factors may provide important context about the paths along which an individual arrived at their current state. This provides important information for interventional decision-making for those with suicidal ideation. Similarly, understanding proximal and distal factors and their effects on suicidal ideation provides information for prevention measures and their relative timings.

For example, the influence of experiencing a traumatic event on lifetime (Zatti et al., 2017) and youth (Barbosa et al., 2014; Hink et al., 2022) STBs is well known. However, the processes by which trauma influences STBs along with other clinical and psychosocial factors is still debated (Hickie et al., 2024). Our analysis shows that a traumatic event influences STBs indirectly through a complex array of clinical and psychosocial factors. It has proximal effects on insomnia, abnormal perceptions, perceived negative social connection, which further influence other factors that then effect suicidal ideation. This cascade of influence provides suggestions for interventional and preventative targets. Preventing a traumatic event is important, but even with knowledge of an individual experiencing a traumatic event, the next factors along the path are likely to be good interventional and preventative targets.

We also show that psychosis-like experiences are a proximal factor of suicidal ideation. This is likely driven by hearing voices and a lack of control of thoughts as shown in the undirected PGM analysis. This is consistent with other evidence linking psychosis to suicidal thoughts, which includes an analysis using undirected PGMs, that showed suicidal thoughts dependent on perceptual anomalies, such as hearing voices and sounds, as well as bizarre experiences (Núñez et al., 2018). The dependency of suicidal ideation on psychosis suggests that theoretical models should include these factors. Those models typically cover depressive and social factors along with extra conditions relating to means and tolerance to a suicide attempt. Such models may need to be improved upon by including psychosis-like experiences.

The theoretical literature on pathways to STBs suggests that other factors may be of importance. Factors that have been proposed include thwarted belongingness, burdensomeness, psychological pain, and hopelessness leading to suicidal ideation. A suicide attempt typically requires suicidal ideation along with a lowered tolerance to and a means of engaging in suicide attempts. We don’t probe all these factors, although some do relate to our factors.

Burdensomeness and thwarted belongness are considered important within the theoretical literature (Van Orden, Witte, Cukrowicz, Braithwaite, Selby, & Joiner, 2010) and are often measured using the interpersonal needs questionnaire (Van Orden et al., 2008). Burdensomeness is partially related to self-perception (although it misses the interpersonal concepts relating to how others view them), which was found to be a bridge node between depression and suicidal ideation within the undirected PGM analysis, but was not found to be an ancestor of suicidal ideation in the BN analysis. Thwarted belongingness is related to several items relating to social connection and interpersonal functioning, which both had direct influence on suicidal ideation.

Psychological pain is a broad concept that has been put forth as a key determinant of STBs (Ducasse et al., 2018; Mee et al., 2006; Orbach et al., 2003). Psychological pain is typically considered to be a construct that goes beyond depressive or clinical symptoms representing a longer lasting negative deficiency of self (Meerwijk & Weiss, 2011). Such a construct may relate to self-perception within our variables, but is likely to go beyond our variables, and thus would be best assessed using a well-defined psychological pain questionnaire (Blandizzi et al., 2025; Holden et al., 2001; Mee et al., 2011; Orbach et al., 2003). Thus, we suspect we do not appropriately probe psychological pain within this analysis.

Burdensomeness, thwarted belongingness, and psychological pain may be more proximal factors to suicidal ideation than the clinical and psychosocial variables within our data. However, our factors may influence these theoretical factors. For example, depressive symptoms are likely to affect psychological pain, whereas burdensomeness and thwarted belongingness may be affected by interpersonal functioning, including but not limited to social connection and personal functioning, which we probe in this analysis.

Only suicidal ideation was found to be a proximal factor to recent suicide attempts. While suicidal ideation is an important outcome in and of itself, and an important prevention target for suicide attempts (Jobes & Joiner, 2019), many individuals with suicidal ideation do not make a suicide attempt. Thus, to understand the paths to suicide attempts, we will need to include variables that are expected to influence the progression from suicidal ideation to attempts, including capability of suicide (Paashaus et al., 2019) along with relevant decision-making processes (Galasiński & Ziółkowska, 2024; I. Xu et al., 2024).

Suicidal ideation fluctuates over both long (e.g., months, years) and short (e.g., minutes, hours) time scales (Kleiman et al., 2018; Varidel et al., 2024; Wang et al., 2024). Also, around 20-60% decide to make an attempt within short time periods (minutes to hours) in adult samples (Cáceda et al., 2020; Paashaus et al., 2019; I. Xu et al., 2024). STBs also predict and may causally affect clinical and psychosocial factors (Iorfino et al., 2018). This suggests that longitudinal short time-scale data will be important to understand the paths to and from STBs. Dynamic probabilistic graphical models will be an aide to unravel temporal dependencies, and thus longitudinal paths, in such data. Li & Kwok (2023) used undirected dynamic PGMs and Iorfino et al. (2023b) used dynamic BNs to understand the temporal dependencies between multiple clinical and psychosocial variables with suicidal ideation. However, both analyses had long time periods between observations (months up to 1 year), and thus would be improved by including shorter-term data, to further unravel the paths to STBs.

In summary, suicidal ideation can arise from a multitude of mental health and behavioural factors for youths engaged in mental health care. We found that depressed mood, personal functional impairment, poor social connection, and psychosis-like experiences were proximal factors to suicidal ideation within our data set. Whereas experiencing a traumatic event and anxiety were important distal factors on suicidal ideation. This provides a range of potential interventional targets along with relative timing of events that could improve interventions.

## Data Availability

All data produced in the present study are available upon reasonable request to the authors.

## References

Altman, E. G., Hedeker, D., Peterson, J. L., & Davis, J. M. (1997). The altman self-rating Mania scale. Biological Psychiatry, 42(10), 948–955. 10.1016/S0006-3223(96)00548-3

Ammerman, B. A., & Jacobucci, R. (2023). The impact of social connection on near-term suicidal ideation. Psychiatry Research, 326(January), 115338. 10.1016/j.psychres.2023.115338

Australian Institute of Health and Welfare. (2024). Deaths in Australia. https://www.aihw.gov.au/reports/life-expectancy-deaths/deaths-in-australia/contents/leading-causes-of-death

Bakken, V., Lydersen, S., Skokauskas, N., Sund, A. M., & Kaasbøll, J. (2025). Protective factors for suicidal ideation and suicide attempts in adolescence: a longitudinal population-based cohort study examining sex differences. BMC Psychiatry, 25(1), 106. 10.1186/s12888-025-06552-6

Barbosa, L. P., Quevedo, L., da Silva, G. D. G., Jansen, K., Pinheiro, R. T., Branco, J. Ô., Lara, D., Oses, J., & da Silva, R. A. (2014). Childhood trauma and suicide risk in a sample of young individuals aged 14-35 years in southern Brazil. Child Abuse and Neglect, 38(7), 1191–1196. 10.1016/j.chiabu.2014.02.008

Bilsen, J. (2018). Suicide and Youth: Risk Factors. In Frontiers in Psychiatry (Vol. 9). Frontiers Media S.A. 10.3389/fpsyt.2018.00540

Biswas, T., Scott, J. G., Munir, K., Renzaho, A. M. N., Rawal, L. B., Baxter, J., & Mamun, A. A. (2020). Global variation in the prevalence of suicidal ideation, anxiety and their correlates among adolescents: A population based study of 82 countries. EClinicalMedicine, 24. 10.1016/j.eclinm.2020.100395

Blandizzi, C., Carlucci, L., Balsamo, M., Contardi, A., Bungaro, N., Erbuto, D., Pompili, M., & Innamorati, M. (2025). Measuring psychache as a suicide risk variable: A Mokken analysis of the Holden’s Psychache Scale. Journal of Affective Disorders, 369, 80–86. 10.1016/j.jad.2024.09.160

Borsboom, D., Deserno, M. K., Rhemtulla, M., Epskamp, S., Fried, E. I., McNally, R. J., Robinaugh, D. J., Perugini, M., Dalege, J., Costantini, G., Isvoranu, A. M., Wysocki, A. C., van Borkulo, C. D., van Bork, R., & Waldorp, L. J. (2021). Network analysis of multivariate data in psychological science. In Nature Reviews Methods Primers (Vol. 1, Issue 1). Springer Nature. 10.1038/s43586-021-00055-w

Bryan, C. J., Butner, J. E., May, A. M., Rugo, K. F., Harris, J. A., Oakey, D. N., Rozek, D. C., & Bryan, A. B. O. (2020). Nonlinear change processes and the emergence of suicidal behavior: A conceptual model based on the fluid vulnerability theory of suicide. New Ideas in Psychology, 57(March 2019), 100758. 10.1016/j.newideapsych.2019.100758

Buysse, D. J., Reynolds, C. F., Monk, T. H., Berman, S. R., & Kupfer, D. J. (1989). The Pittsburgh Sleep Quality Index: a new instrument for psychiatric practice and research. Psychiatry Res. 1989;28:193–213. *Psychiatry Research*, *28*(2), 193–213. 10.1016/0165-1781(89)90047-4

Cáceda, R., Carbajal, J. M., Salomon, R. M., Moore, J. E., Perlman, G., Padala, P. R., Hasan, A., & Delgado, P. L. (2020). Slower perception of time in depressed and suicidal patients. European Neuropsychopharmacology, 40, 4–16. 10.1016/j.euroneuro.2020.09.004

Capon, W., Hickie, I. B., McKenna, S., Varidel, M., Richards, M., LaMonica, H. M., Rock, D., Scott, E. M., & Iorfino, F. (2023). Characterising variability in youth mental health service populations: A detailed and scalable approach using digital technology. Australasian Psychiatry, 31(3), 295–301. 10.1177/10398562231167681

Christiansen, E., Goldney, R. D., Beautrai, A. L., & Agerbo, E. (2011). Youth suicide attempts and the dose-response relationship to parental risk factors: A population-based study. Psychological Medicine, 41(2), 313–319. 10.1017/S0033291710000747

Chu, C., Buchman-Schmitt, J. M., Stanley, I. H., Hom, M. A., Tucker, R. P., Hagan, C. R., Rogers, M. L., Podlogar, M. C., Chiurliza, B., Ringer, F. B., Michaels, M. S., Patros, C. H. G., & Joiner, T. E. (2017). The interpersonal theory of suicide: A systematic review and meta-analysis of a decade of cross-national research. Psychological Bulletin, 143(12), 1313–1345. 10.1037/bul0000123

Consoli, A., Peyre, H., Speranza, M., Hassler, C., Falissard, B., Touchette, E., Cohen, D., Moro, M. R., & Révah-Lévy, A. (2013). Suicidal behaviors in depressed adolescents: Role of perceived relationships in the family. Child and Adolescent Psychiatry and Mental Health, 7(1). 10.1186/1753-2000-7-8

Dablander, F., & Hinne, M. (2019). Node centrality measures are a poor substitute for causal inference. Scientific Reports, 9(1), 1–13. 10.1038/s41598-019-43033-9

De Beurs, D., Fried, E. I., Wetherall, K., Cleare, S., O’ Connor, D. B., Ferguson, E., O’Carroll, R. E., & O’ Connor, R. C. (2019). Exploring the psychology of suicidal ideation: A theory driven network analysis. Behaviour Research and Therapy, 120, 103419. 10.1016/j.brat.2019.103419

Delgadillo, J., Budimir, S., Barkham, M., Humer, E., Pieh, C., & Probst, T. (2023). A Bayesian network analysis of psychosocial risk and protective factors for suicidal ideation. Frontiers in Public Health, 11(3). 10.3389/fpubh.2023.1010264

Ducasse, D., Holden, R. R., Boyer, L., Artéro, S., Raffaella, Guillaume, S., Courtet, P., & Olié, E. (2018). Psychological pain in suicidality: A meta-analysis. Journal of Clinical Psychiatry, 79(3). 10.4088/JCP.16r10732

Franklin, J. C., Ribeiro, J. D., Fox, K. R., Bentley, K. H., Kleiman, E. M., Huang, X., Musacchio, K. M., Jaroszewski, A. C., Chang, B. P., & Nock, M. K. (2017). Risk factors for suicidal thoughts and behaviors: A meta-analysis of 50 years of research. Psychological Bulletin, 143(2), 187–232. 10.1037/bul0000084

Galasiński, D., & Ziółkowska, J. (2024). The end of ambivalence. A narrative perspective on ambivalence in the suicidal process. Suicide and Life-Threatening Behavior. 10.1111/sltb.13101

Gelman, A., Carlin, J. B., Stern, H. S., Dunson, D. B., Vehtari, A., & Rubin, D. B. (2013). Bayesian Data Analysis (3rd Ed.). Chapman and Hall/CRC. 10.1201/9780429258411

Goodrich, B., Gabry, J., Ali, A., & Brilleman, S. (2024). rstanarm: Bayesian applied regression modeling via Stan (R package version 2.32.1). https://mc-stan.org/rstanarm

Han, M. A., Kim, K. S., Ryu, S. Y., Kang, M. G., & Park, J. (2009). Associations between smoking and alcohol drinking and suicidal behavior in Korean adolescents: Korea Youth Behavioral Risk Factor Surveillance, 2006. Preventive Medicine, 49(2–3), 248–252. 10.1016/j.ypmed.2009.06.014

Heapy, C., Haddock, G., & Pratt, D. (2024). The Relationship Between Social Problem-Solving and Suicidal Ideation and Behavior in Adults: A Systematic Review and Meta-Analysis. Clinical Psychology: Science and Practice, 31(4), 419–432. 10.1037/cps0000195

Hickie, I. B., McFarlane, A. C., Ospina-Pinillos, L., Chanen, A. M., Medland, S. E., & Crouse, J. J. (2024). To what extent are depressive or other mood disorders a consequence of earlier traumatic experiences? Research Directions: Depression, 1. 10.1017/dep.2023.16

Hickie, I. B., Scott, E. M., Cross, S. P., Iorfino, F., Davenport, T. A., Guastella, A. J., Naismith, S. L., Carpenter, J. S., Rohleder, C., Crouse, J. J., Hermens, D. F., Koethe, D., Markus Leweke, F., Tickell, A. M., Sawrikar, V., & Scott, J. (2019). Right care, first time: a highly personalised and measurement-based care model to manage youth mental health. Medical Journal of Australia, 211(S9). 10.5694/mja2.50383

Hink, A. B., Killings, X., Bhatt, A., Ridings, L. E., & Andrews, A. L. (2022). Adolescent Suicide—Understanding Unique Risks and Opportunities for Trauma Centers to Recognize, Intervene, and Prevent a Leading Cause of Death. In Current Trauma Reports (Vol. 8, Issue 2, pp. 41–53). Springer Science and Business Media Deutschland GmbH. 10.1007/s40719-022-00223-7

Holden, R. R., Mehta, K., Cunningham, E. J., & Mcleod, L. D. (2001). Development and Preliminary Validation of a Scale of Psychache. Canadian Journal of Behavioural Science/Revue Canadienne Des Sciences Du Comportement, 4(33), 224. 10.1037/h0087144

Holman, M. S., & Williams, M. N. (2020). Suicide Risk and Protective Factors: A Network Approach. Archives of Suicide Research, 1–18. 10.1080/13811118.2020.1774454

Iorfino, F., Cross, S. P., Davenport, T., Carpenter, J. S., Scott, E., Shiran, S., & Hickie, I. B. (2019). A Digital Platform Designed for Youth Mental Health Services to Deliver Personalized and Measurement-Based Care. Frontiers in Psychiatry, 10(August), 1–9. 10.3389/fpsyt.2019.00595

Iorfino, F., F. Hermens, D., Cross, S. P. M., Zmicerevska, N., Nichles, A., Groot, J., Guastella, A. J., Scott, E. M., & B. Hickie, I. (2018). Prior suicide attempts predict worse clinical and functional outcomes in young people attending a mental health service. Journal of Affective Disorders, 238(June), 563–569. 10.1016/j.jad.2018.06.032

Iorfino, F., Varidel, M., Marchant, R., Cripps, S., Crouse, J., Prodan, A., Oliveria, R., Carpenter, J. S., Hermens, D. F., Guastella, A., Scott, E., Shah, J., Merikangas, K., Scott, J., & Hickie, I. B. (2023a). The temporal dependencies between social, emotional and physical health factors in young people receiving mental healthcare: a dynamic Bayesian network analysis. Epidemiology and Psychiatric Sciences, 32, e56. 10.1017/S2045796023000616

Iorfino, F., Varidel, M., Marchant, R., Cripps, S., Crouse, J., Prodan, A., Oliveria, R., Carpenter, J. S., Hermens, D. F., Guastella, A., Scott, E., Shah, J., Merikangas, K., Scott, J., & Hickie, I. B. (2023b). The temporal dependencies between social, emotional and physical health factors in young people receiving mental healthcare: a dynamic Bayesian network analysis. Epidemiology and Psychiatric Sciences, 32. 10.1017/S2045796023000616

Ising, H. K., Veling, W., Loewy, R. L., Rietveld, M. W., Rietdijk, J., Dragt, S., Klaassen, R. M. C., Nieman, D. H., Wunderink, L., Linszen, D. H., & Van Der Gaag, M. (2012). The validity of the 16-item version of the prodromal questionnaire (PQ-16) to screen for ultra high risk of developing psychosis in the general help-seeking population. Schizophrenia Bulletin, 38(6), 1288–1296. 10.1093/schbul/sbs068

Jeon, M. E., Rogers, M. L., Udupa, N., & Joiner, T. E. (2024). Suicidal Ideation and Attempts and Hyperarousal in Military Personnel and Veterans: Network Analysis Reveals Roles of Anxiety Sensitivity and Insomnia. Psychological Trauma: Theory, Research, Practice, and Policy. 10.1037/tra0001685

Jobes, D. A., & Joiner, T. E. (2019). Reflections on Suicidal Ideation. Crisis, 40(4), 227–230. 10.1027/0227-5910/a000615

Kearns, J. C., Coppersmith, D. D. L., Santee, A. C., Insel, C., Pigeon, W. R., & Glenn, C. R. (2020). Sleep problems and suicide risk in youth: A systematic review, developmental framework, and implications for hospital treatment. General Hospital Psychiatry, 63, 141–151. 10.1016/j.genhosppsych.2018.09.011

Kleiman, E. M., Turner, B. J., Fedor, S., Beale, E. E., Picard, R. W., Huffman, J. C., & Nock, M. K. (2018). Digital phenotyping of suicidal thoughts. Depression and Anxiety, 35(7), 601–608. 10.1002/da.22730

Klonsky, E. D., & May, A. M. (2015). The Three-Step Theory (3ST): A New Theory of Suicide Rooted in the “Ideation-to-Action” Framework. International Journal of Cognitive Therapy, 8(2), 114–129.

Klonsky, E. D., May, A. M., & Saffer, B. Y. (2016). Suicide, Suicide Attempts, and Suicidal Ideation. Annual Review of Clinical Psychology, 12, 307–330. 10.1146/annurev-clinpsy-021815-093204

Knipe, D., Padmanathan, P., Newton-Howes, G., Chan, L. F., & Kapur, N. (2022). Suicide and self-harm. The Lancet, 399(10338), 1903–1916. 10.1016/S0140-6736(22)00173-8

Koller, D., & Friedman, N. (2009). Probabilistic Graphical Models: Principles and Techniques. MIT press.

Kuipers, J., & Moffa, G. (2017). Partition MCMC for Inference on Acyclic Digraphs. Journal of the American Statistical Association, 112(517), 282–299. 10.1080/01621459.2015.1133426

Kuipers, J., & Moffa, G. (2022). Bestie: Bayesian Estimation of Intervention Effects (v0.1.5). 10.32614/CRAN.package.Bestie

Li, Y., & Kwok, S. Y. C. L. (2023). A Longitudinal Network Analysis of the Interactions of Risk and Protective Factors for Suicidal Potential in Early Adolescents. Journal of Youth and Adolescence, 52(2), 306–318. 10.1007/s10964-022-01698-y

Maathuis, M., Drton, M., Lauritzen, S., & Wainwright, M. (2018). Handbook of Graphical Models. CRC Press. https://www.crcpress.com/go/handbooks

Makowski, D., Ben-Shachar, M., & Lüdecke, D. (2019). bayestestR: Describing Effects and their Uncertainty, Existence and Significance within the Bayesian Framework. Journal of Open Source Software, 4(40), 1541. 10.21105/joss.01541

May, A. M., & Klonsky, E. D. (2016). What Distinguishes Suicide Attempters From Suicide Ideators? A Meta-Analysis of Potential Factors. Clinical Psychology: Science and Practice, 23(1), 5–20. 10.1111/cpsp.12136

McGorry, P. D., Mei, C., Dalal, N., Alvarez-Jimenez, M., Blakemore, S. J., Browne, V., Dooley, B., Hickie, I. B., Jones, P. B., McDaid, D., Mihalopoulos, C., Wood, S. J., El Azzouzi, F. A., Fazio, J., Gow, E., Hanjabam, S., Hayes, A., Morris, A., Pang, E., … Killackey, E. (2024). The Lancet Psychiatry Commission on youth mental health. The Lancet Psychiatry, 11(9), 731–774. 10.1016/S2215-0366(24)00163-9

McGorry, P. D., Tanti, C., Stokes, R., Hickie, I. B., Carnell, K., Littlefield, L. K., & Moran, J. (2007). headspace: Australia’s National Youth Mental Health Foundation--where young minds come first. The Medical Journal of Australia, *187*(7 Suppl). 10.5694/j.1326-5377.2007.tb01342.x

Mee, S., Bunney, B. G., Bunney, W. E., Hetrick, W., Potkin, S. G., & Reist, C. (2011). Assessment of psychological pain in major depressive episodes. Journal of Psychiatric Research, 45(11), 1504–1510. 10.1016/j.jpsychires.2011.06.011

Mee, S., Bunney, B. G., Reist, C., Potkin, S. G., & Bunney, W. E. (2006). Psychological pain: A review of evidence. In Journal of Psychiatric Research (Vol. 40, Issue 8, pp. 680–690). 10.1016/j.jpsychires.2006.03.003

Meerwijk, E. L., & Weiss, S. J. (2011). Toward a unifying definition of psychological pain. Journal of Loss and Trauma, 16(5), 402–412. 10.1080/15325024.2011.572044

Mohammadi, R., & Wit, E. C. (2019). BDgraph: An R package for Bayesian structure learning in graphical models. Journal of Statistical Software, 89(1). 10.18637/jss.v089.i03

Moran, P., Chandler, A., Dudgeon, P., Kirtley, O. J., Knipe, D., Pirkis, J., Sinyor, M., Allister, R., Ansloos, J., Ball, M. A., Chan, L. F., Darwin, L., Derry, K. L., Hawton, K., Heney, V., Hetrick, S., Li, A., Machado, D. B., McAllister, E., … Christensen, H. (2024). The Lancet Commission on self-harm. The Lancet, 404(10461), 1445–1492. 10.1016/S0140-6736(24)01121-8

Mundt, J. C., Marks, I. M., Shear, M. K., & Greist, J. H. (2002). The Work and Social Adjustment Scale: A simple measure of impairment in functioning. British Journal of Psychiatry, 180(5), 461–464. 10.1192/bjp.180.5.461

Nock, M. K., Borges, G., Bromet, E. J., Alonso, J., Angermeyer, M., Beautrais, A., Bruffaerts, R., Wai, T. C., De Girolamo, G., Gluzman, S., De Graaf, R., Gureje, O., Haro, J. M., Huang, Y., Karam, E., Kessler, R. C., Lepine, J. P., Levinson, D., Medina-Mora, M. E.,… Williams, D. (2008). Cross-national prevalence and risk factors for suicidal ideation, plans and attempts. British Journal of Psychiatry, 192(2), 98–105. 10.1192/bjp.bp.107.040113

Nock, M. K., Green, J. G., Hwang, I., McLaughlin, K. A., Sampson, N. A., Zaslavsky, A. M., & Kessler, R. C. (2013). Prevalence, correlates, and treatment of lifetime suicidal behavior among adolescents: Results from the national comorbidity survey replication adolescent supplement. JAMA Psychiatry, 70(3), 300–310. 10.1001/2013.jamapsychiatry.55

Norman, S. B., Cissell, S. H., Means-Christensen, A. J., & Stein, M. B. (2006). Development and Validation of an Overall Anxiety Severity and Impairment Scale (OASIS). Depression and Anxiety, 23(4), 245–249. 10.1002/da.20182

Núñez, D., Fresno, A., van Borkulo, C. D., Courtet, P., Arias, V., Garrido, V., & Wigman, J. T. W. (2018). Examining relationships between psychotic experiences and suicidal ideation in adolescents using a network approach. Schizophrenia Research, 201, 54–61. 10.1016/j.schres.2018.05.020

O’Connor, R. C., & Kirtley, O. J. (2018). The integrated motivational-volitional model of suicidal behaviour. Philosophical Transactions of the Royal Society B: Biological Sciences, 373(1754). 10.1098/rstb.2017.0268

O’connor, R. C., & Nock, M. K. (2014). The psychology of suicidal behaviour. 73. 10.1016/S2215-0366(14)70222-6

Orbach, I., Mikulincer, M., Gilboa-Schechtman, E., & Sirota, P. (2003). Mental Pain and Its Relationship to Suicidality and Life Meaning. Suicide and Life-Threatening Behavior, 33(3), 231–241. 10.1521/suli.33.3.231.23213

Orri, M., Scardera, S., Perret, L. C., Bolanis, D., Temcheff, C., Séguin, J. R., Boivin, M., Turecki, G., Tremblay, R. E., Côté, S. M., & Geoffroy, M.-C. (2020). Mental Health Problems and Risk of Suicidal Ideation and Attempts in Adolescents. Pediatrics, 146. http://publications.aap.org/pediatrics/article-pdf/146/1/e20193823/1079539/peds_20193823.pdf

Paashaus, L., Forkmann, T., Glaesmer, H., Juckel, G., Rath, D., Schönfelder, A., Engel, P., & Teismann, T. (2019). Do suicide attempters and suicide ideators differ in capability for suicide? Psychiatry Research, 275, 304–309. 10.1016/j.psychres.2019.03.038

Pearl, J., Glymour, M., & Jewell, N. P. (2016). Causal Inference in Statistics: A Primer. John Wiley & Sons, Ltd.

Posner, K., Brown, G. K., & Stanley, B. (2011). The Columbia–Suicide Severity Rating Scale: Initial Validity and Internal Consistency Findings From Three Multisite Studies With Adolescents and Adults. American Journal of Psychiatry, 168(12), 1267–1277. 10.1176/appi.ajp.2011.10111704

R Core Team. (2024). R: A Language and Environment for Statistical Computing. R Foundation for Statistical Computing. https://www.r-project.org/

Roenneberg, T., Wirz-Justice, A., & Merrow, M. (2003). Life between clocks: Daily temporal patterns of human chronotypes. Journal of Biological Rhythms, 18(1), 80–90. 10.1177/0748730402239679

Rugo-Cook, K. F., Kerig, P. K., Crowell, S. E., & Bryan, C. J. (2021). Fluid vulnerability theory as a framework for understanding the association between posttraumatic stress disorder and suicide: A narrative review. Journal of Traumatic Stress, 34(6), 1080–1098. 10.1002/jts.22782

Rush, A. J., Trivedi, M. H., Ibrahim, H. M., Carmody, T. J., Arnow, B., Klein, D. N., Markowitz, J. C., Ninan, P. T., Kornstein, S., Manber, R., Thase, M. E., Kocsis, J. H., & Keller, M. B. (2003). The 16-item Quick Inventory of Depressive Symptomatology (QIDS), clinician rating (QIDS-C), and self-report (QIDS-SR): A psychometric evaluation in patients with chronic major depression. Biological Psychiatry, 54(5), 573–583. 10.1016/S0006-3223(02)01866-8

Ryan, O., Bringmann, L. F., & Schuurman, N. K. (2022). The Challenge of Generating Causal Hypotheses Using Network Models. Structural Equation Modeling, 29(6), 953–970. 10.1080/10705511.2022.2056039

Schuster, T. L., Kessler, R. C., & Aseltine, R. H. (1990). Supportive interactions, negative interactions, and depressed mood. American Journal of Community Psychology, 18(3), 423–438. 10.1007/BF00938116

Scutari, M. (2010). Learning Bayesian Networks with the bnlearn R package. Journal of Statistical Software, 35(3), 1–22.

Scutari, M., Graafland, C. E., & Gutiérrez, J. M. (2019). Who learns better Bayesian network structures: Accuracy and speed of structure learning algorithms. International Journal of Approximate Reasoning, 115, 235–253. 10.1016/j.ijar.2019.10.003

Skinner, A., Osgood, N. D., Occhipinti, J. A., Song, Y. J. C., & Hickie, I. B. (2023). Unemployment and underemployment are causes of suicide. Science Advances, 9(28), eadg3758. 10.1126/sciadv.adg3758

Suter, P., Kuipers, J., Moffa, G., & Beerenwinkel, N. (2023). Bayesian Structure Learning and Sampling of Bayesian Networks with the R Package BiDAG. Journal of Statistical Software, 105(9), 1–31. 10.18637/jss.v105.i09

Uddin, R., Burton, N. W., Maple, M., Khan, S. R., & Khan, A. (2019). Suicidal ideation, suicide planning, and suicide attempts among adolescents in 59 low-income and middle-income countries: a population-based study. The Lancet Child & Adolescent Health, 3, 223–233. 10.1016/S2352-4642(18)30403-6

Van Meter, A. R., Knowles, E. A., & Mintz, E. H. (2023). Systematic Review and Meta-analysis: International Prevalence of Suicidal Ideation and Attempt in Youth. Journal of the American Academy of Child & Adolescent Psychiatry, 9(62), 973–986. 10.1016/j.jaac.2022.07.867

Van Orden, K. A., Witte, T. K., Cukrowicz, K. C., Braithwaite, S. R., Selby, E. A., & Joiner, T. E. (2010). The Interpersonal Theory of Suicide. Psychological Review, 117(2), 575–600. 10.1037/a0018697

Van Orden, K. A., Witte, T. K., Cukrowicz, K. C., Braithwaite, S. R., Selby, E. A., Joiner, T. E., & Jr. (2010). The Interpersonal Theory of Suicide. Psychological Review, 117(2), 575–600. 10.1037/a0018697.The

Van Orden, K. A., Witte, T. K., Gordon, K. H., Bender, T. W., & Joiner, T. E. (2008). Suicidal Desire and the Capability for Suicide: Tests of the Interpersonal-Psychological Theory of Suicidal Behavior Among Adults. Journal of Consulting and Clinical Psychology, 76(1), 72–83. 10.1037/0022-006X.76.1.72

Van Spijker, B. A. J., Batterham, P. J., Calear, A. L., Farrer, L., Christensen, H., Reynolds, J., & Kerkhof, A. J. F. M. (2014). The Suicidal Ideation Attributes Scale (SIDAS): Community-based validation study of a new scale for the measurement of suicidal ideation. Suicide and Life-Threatening Behavior, 44(4), 408–419. 10.1111/sltb.12084

Varidel, M. (2024). CIA: Learn and Apply Directed Acyclic Graphs for Causal Inference (1.0.0). Zenodo. 10.5281/zenodo.14176795

Varidel, M., Hickie, I., Prodan, A., Skinner, A., Marchant, R., & … (2024). Dynamic learning of individual-level suicidal ideation trajectories to enhance mental health care. Npj Mental Health Research, 3(1), 26. 10.1038/s44184-024-00071-0

Vehtari, A., Gelman, A., Simpson, D., Carpenter, B., & Burkner, P. C. (2021). Rank-Normalization, Folding, and Localization: An Improved R-hat for Assessing Convergence of MCMC (with Discussion). Bayesian Analysis, 16(2), 667–718. 10.1214/20-BA1221

Vernon, M. K., Dugar, A., Revicki, D., Treglia, M., & Buysse, D. (2010). Measurement of non-restorative sleep in insomnia: A review of the literature. Sleep Medicine Reviews, 14(3), 205–212. 10.1016/j.smrv.2009.10.002

Wang, S. B., Van Genugten, R. DI, Yacoby, Y., Pan, W., Bentley, K. H., Bird, S. A., Buonopane, R. J., Christie, A., Daniel, M., DeMarco, D., Haim, A., Follet, L., Fortgang, R. G., Kelly, F., Kleiman, E. M., Millner, A. J., Obi-Obasi, O., Onnela, J., Ramlal, N., … Paulson, J. A. (2024). Idiographic Prediction of Suicidal Thoughts: Building Personalized Machine Learning Models with Real-Time Monitoring Data. Nature Mental Health, 2(November). 10.1038/s44220-024-00335-w

World Health Organization. (2021). Suicide worldwide in 2019: Global Health Estimates. In World Health Organization,Geneva.

Xu, I., Millner, A. J., Fortgang, R. G., & Nock, M. K. (2024). Suicide decision-making: Differences in proximal considerations between individuals who aborted and attempted suicide. Suicide and Life-Threatening Behavior. 10.1111/sltb.13127

Xu, Y. E., Barron, D. A., Sudol, K., Zisook, S., & Oquendo, M. A. (2023). Suicidal behavior across a broad range of psychiatric disorders. Molecular Psychiatry, December 2022, 1–47. 10.1038/s41380-022-01935-7

Zatti, C., Rosa, V., Barros, A., Valdivia, L., Calegaro, V. C., Freitas, L. H., Ceresér, K. M. M., Rocha, N. S. da, Bastos, A. G., & Schuch, F. B. (2017). Childhood trauma and suicide attempt: A meta-analysis of longitudinal studies from the last decade. Psychiatry Research, 256, 353–358. 10.1016/j.psychres.2017.06.082

Zhang, J., Lamers, F., Hickie, I. B., He, J. P., Feig, E., & Merikangas, K. R. (2013). Differentiating nonrestorative sleep from nocturnal insomnia symptoms: Demographic, clinical, inflammatory, and functional correlates. Sleep, 36(5), 671–679. 10.5665/sleep.2624

