## Supplementary Material for "Pathways to suicidal ideation for young people engaged in mental health care"

### 1. Innowell questionnaire

The following is adapted from Capon, et al.(Capon et al., 2023).

**Overall health**
*EQ-5D-Y* – The EQ-5D-Y (Youth) is a questionnaire that measures the generic health-related quality of life. The Innowell platform only uses a single question related to an individual's health, which asks "How good is your health TODAY? Please respond using a number between 0 (the worst health you can imagine) and 100 (The best health you can imagine)." The individual is given a 100-point scale from which they can select their answer.

**Overall mental health**
*CGI-S* – The Clinical Global Impressions – Severity scale measures an individual's current mental health. It asks a single question; "How would you rate your mental health at this time?". The individual has a selection of 7 items that range from "Normal, not at all unwell" (1) to "Extremely unwell" (7).

**Suicidality**
*SIDAS* – The Suicidal Ideation Attributes Scale (SIDAS) is a five-item scale assessing suicidal ideation over the past month. The scale assesses frequency, controllability, closeness to attempt, distress and interference with daily activities on a 10-point Likert scale (ranging from 0 ("never") to 10 ("always")). A total score of zero corresponds to "no current ideation", a score of one to 20 corresponds to "low current suicidal ideation", and a score of 21 to 50 corresponds to "high current suicidal ideation". The scale has strong internal reliability (Cronbach $\alpha = 0.91$) (Van Spijker et al., 2014).

*C-SSRS* – The Columbia-Suicide Severity Rating Scale (C-SSRS) aims to measure suicidal thoughts and behaviours in the past month and entire lifetime (Posner et al., 2011). The questionnaire assesses four constructs: severity of ideation, intensity of ideation, suicidal behaviours, and lethality.

**Distress**
*K-10* – The Kessler-10 (K-10) measures psychological distress over the past 4 weeks(Kessler et al., 2002). It is a well-validated measure widely used in adult and adolescent populations in both clinical and community settings. The scale consists of 10 items with five multiple choice options ranging from "none of the time" (1) to "all of the time" (5). Total scores range from 10-50. A total score of under 20 indicates 'likely no distress', 20-24 indicates "likely mild mental disorder", 25-29 indicates "likely moderate mental disorder", over 30 indicates 'likely severe mental disorder' (Andrews & Slade, 2001). The K-10 has moderate reliability (kappa scores 0.42-0.74).

**Psychosis-like experiences**
*PQ-16* – The Prodromal Questionnaire (PQ-16) is a self-report measure used to screen for individuals at risk of psychosis and select individuals for interview of psychosis risk (Ising et al., 2012). It was adapted from the 92-item Prodromal Questionnaire, including nine items from the perceptual abnormalities subscale, five items from the unusual thought content/delusional ideas subscale, and two items from the negative symptoms subscale (Howie et al., 2022). Questions require "True" or "False" answers to statements describing feelings, experiences, or symptoms of psychosis. In the event of a "True" response, the participant is required to then interpret the perceived distress of that on a Likert scale (0-3, with 3 being severe distress). A

cut-off score of 6 or more on the symptom-scale has been shown to detect at-risk mental states with 87% specificity and sensitivity (Ising et al., 2012).

**Mania-like experiences**
*ASRM* – The Altman Self-Rating Mania Scale (ASRM) is a 5-item scale used to assess the presence and/or severity of manic-like symptoms during the past 7 days. Each item is scored on a 5-point scale (0-4), with total scores ranging from 0-20 and higher scores indicating greater severity. A score greater than 5 indicates a high likelihood of a manic or hypomanic condition, as per 86% sensitivity and 87% sensitivity (Altman et al., 1997).

**Functioning**
*Youth not in education or employment (NEET)* – Engagement with either employment, education, or training was based on questions from the Organisation of Economic Co-operation and Development (OECD), and Census of Population and Housing, Australian Bureau of Statistics (ABS)(OECD, 2023). These four multiple choice questions were: 1) Are you currently engaged in education or study (e.g. school, TAFE or university) on a regular basis?; 2) Are you currently engaged in paid employment or work on a regular basis?; 3) Are you currently engaged in voluntary work through an organisation or group on a regular basis?; and 4) Are you currently providing unpaid care, help or assistance to family members or others because of a disability, a long term illness or problems related to old age on a regular basis? Individuals not involved in employment, education, or training were classified as NEET.

*WSAS* – The Work and Social Adjustment Scale (WSAS) is a brief and reliable measure of work and social adjustment. The questions aim to assess whether an individual is currently impaired or unable to perform day-to-day tasks, due to their mental health. The scale consists of five items that require the individual to rate between 0-8 ("not at all" to "very severely"), based on their agreement with the statement. A maximum total score is achieved by summing all 5 items. A total score greater than 20 suggests moderately severe psychopathology, 10-20 suggests significant functional impairment with less severe symptomatology, and scores under 10 are associated with subclinical populations. The scale has a test-retest correlation of 0.73 and has correlations of 0.76 for severity of depression and 0.61 for obsessive-compulsive disorder symptoms(Mundt et al., 2002).

*SOFAS* – The Social and Occupational Assessment Scale is a single question that asks an individual to rate their social and occupational functioning (Goldman et al., 1992). The question on the Innowell platform asks, "Thinking about your ability to participate in everyday social, school (including university/college) or work activities, how much of your "usual activities have been limited?". The individual is then allowed to select 7 options that range from "Only everyday problems or concerns (e.g., mild anxiety before an exam or an occasional argument with family members)" (1) to "Inability to participate in almost all areas of life (e.g., stay in bed all day, or no job, home or friends or think about harming yourself)" (7).

**Alcohol, tobacco, cannabis use**
*ASSIST* – The Alcohol, Smoking and Substance Involvement Screening Test (ASSIST) is an 8-item questionnaire that aims to detect substance use-related problems. It screens for use of tobacco, alcohol, cannabis, cocaine, amphetamine-type stimulants, sedatives and sleeping pills, hallucinogens, inhalants, opioids, and other drugs. Besides the first question which concerns life-time experiences, each question requires the individual to respond to questions that concern the prior 3-months on either 5- or 3- point Likert scales (Humeniuk et al., 2008).

*AUDIT-C* – The Alcohol Use Disorders Identification Test (AUDIT-C) is a brief measure consisting of three questions related to the frequency of general and binge alcohol consumption over the past year (Babor et al., 2001). Each question has 5 options that increase in frequency or quantity of consumption (scores range from 0-4) (Bush et al., 1998). A total score of 0-3 indicates low-risk drinking, 4-5 moderate risk, and scores greater than five indicates high risk drinking, however, this may not apply if total points come from q1 (i.e., when q2 and q3 =0).

**Social connection**
*SSSS* – The Schuster's Social Support Scale (SSSS) is a 6-item questionnaire that aims to assess the frequency of both supportive and negative interactions with family and friends(Schuster et al., 1990). The first five questions are scored on a 4-point Likert scale (0-3; "never" to "often") with alpha reliability ranging from 0.56-0.75, and the sixth question requires a Yes/No response (Schuster et al., 1990).

**Depression**
*QIDS-SR* – The Quick Inventory of Depressive Symptomatology – Self-report (QIDS-SR) consists of 16-items that aim to assess nine domains of depression during the preceding seven days (Rush et al., 2003). These domains are related to the DSM-IV diagnosis of a major depressive disorder, including: sleep, mood, appetite/weight, concentration/decision making, self-view, suicidal ideation, general interest, energy level, and agitation. Each item is scored on a scale between 0-3 points and scoring instructions determine the total score (which ranges from 0-27) (Brown et al., 2008). Scores greater than 21 indicate very severe depression, 16-20 indicate severe depression, 11-15 indicate moderate depression, 6-10 indicate mild depression, and scores of 5 or lower indicate no depression.

**Anxiety**
*OASIS* – The Overall Anxiety Severity and Impairment Scale aims to assess the severity and impairment of anxiety-related symptoms over the past seven days (Norman et al., 2006). The scale consists of five multiple choice questions with five options that are scored from 0-4, with higher scores indicating greater severity and/or impairment. A cut-off score of 8 has high validity (87%) for identifying anxiety disorders (Campbell-Sills et al., 2009).

**Physical health**
*Height, weight, and waist circumference*
Body mass index (BMI) is calculated by Weight/(Height$^2$) to estimate total body fat in proportion to total body weight. It is used to estimate risk of cardiovascular, metabolic, and other diseases. Waist circumference, however, is a more accurate estimate of visceral fat and more predictive of cardiovascular diseases. For women, a waist circumference of 80-87cm is considered increased risk, and 88+cm is greatly increased. For men, a waist circumference of 94-101cm is considered increased risk, and 102+cm is considered greatly increased risk (Department of Health and Aged Care, 2021).

*IPAQ* – The International Physical Activity Questionnaire (IPAQ) aims to determine the average physical activity of an individual and can be scored on a continuous and/or categorial scale (Craig et al., 2003). First, all activity is calculated in minutes. Second, minutes should be converted to metabolic equivalent of task (MET) minutes (multiply the minutes by the relevant scalar; walking =3.3, moderate activity=4, vigorous activity =8). Third, multiply MET minutes by number of days the activity was performed. For categorical scoring, high = over 3000 MET minutes a week OR over 1500 MET minutes per week with 3 or more days of vigorous exercise; moderate = at least 3 days of vigorous activity or walking of 30 or more mins per day,

OR 5+ days of moderate intensity activity and/or walking (minimum 30 mins per day), OR 5+ days of walking, moderate intensity, or vigorous activity equating to 600+ MET minutes; and low = not meeting high or moderate.

**Sleep-wake cycle**

*PSQI* – The Pittsburgh Sleep Quality Index (PSQI) is a questionnaire that aims to assess sleep quality and disturbances over the past month (Buysse et al., 1989). It consists of nineteen-items that assess seven domains: sleep quality, sleep latency, sleep duration, habitual sleep efficiency, sleep disturbances, use of sleeping medication, and daytime dysfunction. Each of the seven component scores are summed together to form a global score (Buysse et al., 1989).

*MCTQ* – The Munich Chronotype Questionnaire (MCTQ) is a self-report scale that assesses bed- and rise-times, and self-assessment of individual chronotype (Roenneberg et al., 2003). The chronotype options range from extremely early to extremely late and is determined by using the midpoint between onset and offset of sleep.

**Post-traumatic stress**

*PC-PTSD-5* – The Primary Care PTSD Screen (PC-PTSD) is a five-item questionnaire that aims to assess PTSD symptoms which reflect the DSM-V diagnostic criteria (Prins et al., 2016). Each item is scored as either Yes or No (1 or 0), and a maximum total score is 5. A cut-off of 3 is optimally sensitive (reduces false negatives; $\kappa[1]) = 0.93$, standard error = 0.041), yet a cut-off of 4 is optimally efficient (good balance between false positive and negatives; $\kappa[0.5] = 0.63$, standard error = 0.052) (Prins et al., 2016).

**Eating behaviours and body image**

*EDE* – The Eating Disorder Examination Questionnaire (EDE-Q) is based on the eating disorder examination interview and aims to assess eating behaviours and body image disturbance over the past four weeks (Hay et al., 2008). The questionnaire uses a combination of Likert scales and Yes/No options.

### 2. Factors

| Factor | Item |
|---|---|
| Trauma Event | PTSD5_EVENT |
| Insomnia | QIDS_1 |
| | QIDS_2 |
| | QIDS_3 |
| Hypersomnia | QIDS_4 |
| Anxiety | OASIS_1 |
| | OASIS_2 |
| Unrefreshed Sleep | SLEEPWAKE_6 |
| Energy | QIDS_14 |
| Interest | QIDS_13 |
| Psychomotor | QIDS_15 |
| | QIDS_16 |
| Unusual Thoughts | PQ16_2 |
| | PQ16_5 |
| | PQ16_10 |
| | PQ16_11 |
| | PQ16_14 |
| Abnormal Perceptions | PQ16_3 |
| | PQ16_4 |
| | PQ16_6 |
| | PQ16_8 |
| | PQ16_9 |
| | PQ16_12 |
| | PQ16_13 |
| | PQ16_14 |
| | PQ16_15 |
| | PQ16_16 |
| Social Positive | SSSS_1 |
| | SSSS_2 |
| | SSSS_6 |
| Social Negative | SSSS_3 |
| | SSSS_4 |
| | SSSS_5 |

**Table S1.** Mapping of items to factors used within the Bayesian network analysis.

### 3. Bayesian Logistic Regression

We predict two STB outcomes, where we consider the $k$-th outcome for individual $i$ given by $y_{ik}$. The predictors are given by clinical, psychosocial, and demographic characteristics. The $j$-th characteristic for individual $i$ is given by $x_{j,i}$. When conducting the marginal logistic regression analysis, we assume that the probability of $y_{k,i}$ is given by the model,

$$p(y_{k,i}|x_{j,i}) = \text{logit}(\beta_{j,k}x_{j,i} + c_k),$$

where $c_k$ is a constant for the $k$-th characteristic.

For the conditional logistic regression we include all sample characteristics. Thus, we assume that the probability model is given by,

$$p(y_{k,i}|x_{1,i}, x_{2,i}, \dots, x_{10,i}) = \text{logit}\left(\sum_{j=1}^{10} \alpha_{j,k}x_{j,i} + d_k\right),$$

where $d_k$ is a constant for the $k$-th characteristic.

The supplementary results Bayesian logistic regression along with further detail about the distribution of the sample characteristics for a recent suicide attempt and high suicidal ideation are shown in Tables S3-S5 and Tables S6-S8, respectively. For all instances of the regression analysis we ran rstanarm (Goodrich et al., 2024) for 4000 iterations per chain across four chains. We used the default weakly informative priors implemented in rstanarm. We report the mean and standard deviation for $\alpha_{j,k}$ and $\beta_{j,k}$ as the distribution is typically symmetric. Whereas we report the median and highest density interval for the odds-ratio in the main article as the distribution is typically asymmetric. The $\hat{R}$ and effective sample sizes are also reported as convergence statistics (Gelman et al., 2013; Vehtari et al., 2021).

**Table S3.** Categorical clinical and psychosocial characteristics for the sample split by having a recent suicide attempt.

| | Total | Recent Suicide Attempt | No Recent Suicide Attempt |
|---|---|---|---|
| No. (%) | 1020 | 21 (2.1%) | 999 (97.9%) |
| **Depressed mood (QIDS)** | | | |
| Minimal (0-5) | 7 (0.7%) | 0 (0.0%) | 7 (0.7%) |
| Mild (6-10) | 92 (9.0%) | 0 (0.0%) | 92 (9.2%) |
| Moderate (11-15) | 371 (36.4%) | 1 (4.8%) | 370 (37.0%) |
| Severe (16-20) | 409 (40.1%) | 10 (47.6%) | 399 (39.9%) |
| Very severe (21-27) | 141 (13.8%) | 10 (47.6%) | 131 (13.1%) |
| **Psychosis-like experiences (PQ16)** | | | |
| No concern (0-5) | 501 (49.1%) | 5 (23.8%) | 496 (49.6%) |
| Possible concern (6-9) | 303 (29.7%) | 4 (19.0%) | 299 (29.9%) |
| Probable concern (10-16) | 216 (21.2%) | 12 (57.1%) | 204 (20.4%) |
| **Mania-like experiences (ASRM)** | | | |
| No or possible concern (0-6) | 847 (83.0%) | 16 (76.2%) | 831 (83.2%) |
| Probable concern (6-18) | 173 (17.0%) | 5 (23.8%) | 168 (16.8%) |
| **Social and Occupational Functioning (WSAS)** | | | |
| Mild (0-10) | 65 (6.4%) | 1 (4.8%) | 64 (6.4%) |
| Moderate (10-20) | 430 (42.2%) | 4 (19.0%) | 426 (42.6%) |
| Severe (> 20) | 525 (51.5%) | 16 (76.2%) | 509 (51.0%) |
| **Anxiety (OASIS)** | | | |
| Low (0-5) | 128 (12.5%) | 3 (14.3%) | 125 (12.5%) |
| Mild (6-9) | 265 (26.0%) | 1 (4.8%) | 264 (26.4%) |
| Moderate (10-11) | 211 (20.7%) | 2 (9.5%) | 209 (20.9%) |
| Severe (> 12) | 416 (40.8%) | 15 (71.4%) | 401 (40.1%) |
| **Social Connectedness (SSSS)** | | | |
| Good (0-5) | 268 (26.3%) | 3 (14.3%) | 265 (26.5%) |
| Moderate (6-10) | 489 (47.9%) | 8 (38.1%) | 481 (48.1%) |
| Poor (11-18) | 263 (25.8%) | 10 (47.6%) | 253 (25.3%) |
| **Sleep-Wake Cycle** | | | |
| Good | 85 (8.3%) | 1 (4.8%) | 84 (8.4%) |
| Moderate | 341 (33.4%) | 6 (28.6%) | 335 (33.5%) |
| Poor | 594 (58.2%) | 14 (66.7%) | 580 (58.1%) |

**Table S4.** Marginal logistic regression results for each $\beta_{j,1}$ parameter predicting a recent suicide attempt. We present the median, 95% highest-density interval, $\hat{R}$ and effective sample ($N_{\text{eff}}$) size convergence statistics.

| | Mean | Standard Deviation | $\hat{R}$ | $N_{\text{eff}}$ |
|---|---|---|---|---|
| *Demographics* | | | | |
| Female sex at birth | 1.451 | 0.763 | 1.001 | 3078 |
| Age in years, mean | -0.090 | 0.079 | 1.000 | 4515 |
| Urban, No. | -0.272 | 0.536 | 1.001 | 4313 |
| *Traumatic Event* | 1.05 | 0.519 | 1.000 | 3318 |
| *Total Scores* | | | | |
| Suicidal ideation (SIDAS) | 0.193 | 0.028 | 1.001 | 1668 |
| Depressed mood (QIDS-adjusted†) | 0.298 | 0.064 | 1.000 | 2459 |
| Anxiety (OASIS) | 0.158 | 0.062 | 1.001 | 3005 |
| Psychosis-like experiences (PQ16) | 0.194 | 0.054 | 1.000 | 3127 |
| Mania-like experiences (ASRM) | 0.055 | 0.068 | 1.000 | 4549 |
| Functioning (WSAS) | 0.070 | 0.029 | 1.002 | 3546 |
| Social support (SSSS) | 0.117 | 0.061 | 1.001 | 3843 |
| Sleep (BMC Sleep-Wake Scale) | 0.359 | 0.369 | 1.001 | 4634 |

† QIDS-adjusted is calculated as the total QIDS score with the value of question 12, which asks about suicidal ideation, subtracted.

**Table S7.** Conditional logistic regression results for each $\alpha_{j,1}$ parameter predicting a recent suicide attempt. We present the median, 95% highest-density interval, $\hat{R}$ and effective sample size ($N_{\text{eff}}$) convergence statistics.

| | Mean | Standard Deviation | R_hat | $N_{\text{eff}}$ |
|---|---|---|---|---|
| *Demographics* | | | | |
| Female sex at birth | 0.678 | 0.828 | 1.000 | 8183 |
| Age in years, mean | 0.117 | 0.121 | 1.001 | 6985 |
| Urban, No. | -0.224 | 0.675 | 1.000 | 7490 |
| *Traumatic Event* | 0.691 | 0.664 | 1.000 | 8395 |
| *Total Scores* | | | | |
| Suicidal ideation (SIDAS) | 0.232 | 0.036 | 1.000 | 5163 |
| Depressed mood (QIDS-adjusted†) | 0.104 | 0.108 | 1.000 | 6888 |
| Anxiety (OASIS) | -0.044 | 0.078 | 1.000 | 7849 |
| Psychosis-like experiences (PQ16) | 0.101 | 0.077 | 1.000 | 7453 |
| Mania-like experiences (ASRM) | 0.101 | 0.083 | 1.000 | 8162 |
| Functioning (WSAS) | -0.107 | 0.049 | 1.000 | 6813 |
| Social support (SSSS) | -0.047 | 0.077 | 1.000 | 8487 |
| Sleep (BMC Sleep-Wake Scale) | -0.276 | 0.461 | 1.000 | 9277 |

† QIDS-adjusted is calculated as the total QIDS score with the value of question 12, which asks about suicidal ideation, subtracted.

**Table S6.** Categorical clinical and psychosocial characteristics for the sample split by having high (SIDAS ≥ 20) compared to low suicidal ideation.

| | **Total** | **High Suicidal Ideation** | **Low Suicidal Ideation** |
|---|---|---|---|
| No. (%) | 1020 | 321 (31.5%) | 699 (68.5%) |
| **Depressed mood (QIDS)** | | | |
| Minimal (0-5) | 7 (0.7%) | 0 (0%) | 7 (1.0%) |
| Mild (6-10) | 92 (9.0%) | 3 (0.9%) | 89 (12.7%) |
| Moderate (11-15) | 371 (36.4%) | 70 (21.8%) | 301 (43.1%) |
| Severe (16-20) | 409 (40.1%) | 148 (46.1%) | 261 (37.3%) |
| Very severe (21-27) | 141 (13.8%) | 100 (31.2%) | 41 (5.9%) |
| **Psychosis-like experiences (PQ16)** | | | |
| No concern (0-5) | 501 (49.1%) | 120 (37.4%) | 381 (54.5%) |
| Possible concern (6-9) | 303 (29.7%) | 96 (29.9%) | 207 (29.6%) |
| Probable concern (10-16) | 216 (21.2%) | 105 (32.7%) | 111 (15.9%) |
| **Mania-like experiences (ASRM)** | | | |
| No or possible concern (0-6) | 847 (83.0%) | 269 (83.8%) | 578 (82.7%) |
| Probable concern (6-18) | 173 (17.0%) | 52 (16.2%) | 121 (17.3%) |
| **Social and Occupational Functioning (WSAS)** | | | |
| Mild (0-10) | 65 (6.4%) | 6 (1.9%) | 59 (8.4%) |
| Moderate (10-20) | 430 (42.2%) | 81 (25.2%) | 349 (49.9%) |
| Severe (> 20) | 525 (51.5%) | 234 (72.9%) | 291 (41.6%) |
| **Anxiety (OASIS)** | | | |
| Low (0-5) | 128 (12.5%) | 28 (8.7%) | 100 (14.3%) |
| Mild (6-9) | 265 (26.0%) | 55 (17.1%) | 210 (30.0%) |
| Moderate (10-11) | 211 (20.7%) | 63 (19.6%) | 148 (21.2%) |
| Severe (> 12) | 416 (40.8%) | 175 (54.5%) | 241 (34.5%) |
| **Social Connectedness (SSSS)** | | | |
| Good (0-5) | 268 (26.3%) | 55 (17.1%) | 213 (30.5%) |
| Moderate (6-10) | 489 (47.9%) | 145 (45.2%) | 344 (49.2%) |
| Poor (11-18) | 263 (25.8%) | 121 (37.7%) | 142 (20.3%) |
| **BMC Sleep-Wake Cycle** | | | |
| Good | 85 (8.3%) | 16 (5.0%) | 69 (9.9%) |
| Moderate | 341 (33.4%) | 98 (30.5%) | 243 (34.8%) |
| Poor | 594 (58.2%) | 207 (64.5%) | 387 (55.4%) |

**Table S5.** Marginal logistic regression results for each $\beta_{j,2}$ parameter predicting a high suicidal ideation. We present the median, 95% highest-density interval, $\hat{R}$ and effective sample size ($N_{\text{eff}}$) convergence statistics.

| | Mean | Standard Deviation | $\widehat{\boldsymbol{R}}$ | $\boldsymbol{N}_{\text{eff}}$ |
|---|---|---|---|---|
| *Demographics* | | | | |
| Female sex at birth | 0.258 | 0.159 | 1.000 | 6004 |
| Age in years, mean | -0.071 | 0.024 | 1.000 | 5912 |
| Urban, No. | -0.249 | 0.175 | 1.000 | 5807 |
| *Traumatic Event* | 0.396 | 0.139 | 1.000 | 5253 |
| *Total Scores* | | | | |
| Depressed mood (QIDS-adjusted[†]) | 0.227 | 0.022 | 1.000 | 5057 |
| Anxiety (OASIS) | 0.127 | 0.019 | 1.000 | 5111 |
| Psychosis-like experiences (PQ16) | 0.128 | 0.018 | 1.001 | 4794 |
| Mania-like experiences (ASRM) | -0.019 | 0.024 | 1.000 | 5346 |
| Functioning (WSAS) | 0.106 | 0.010 | 1.001 | 4124 |
| Social support (SSSS) | 0.138 | 0.020 | 1.000 | 4980 |
| Sleep (BMC Sleep-Wake Scale) | 0.355 | 0.112 | 1.000 | 5660 |

[†] QIDS-adjusted is calculated as the total QIDS score with the value of question 12, which asks about suicidal ideation, subtracted.

**Table S8.** Conditional logistic regression results for each $\alpha_{j,2}$ parameter predicting high suicidal ideation. We present the mean, standard deviation, $\hat{R}$, and effective sample size convergence ($N_{\text{eff}}$) statistics.

| | Mean | Standard Deviation | R_hat | $\boldsymbol{N}_{\text{eff}}$ |
|---|---|---|---|---|
| *Demographics* | | | | |
| Female sex at birth | 0.072 | 0.174 | 1.000 | 10110 |
| Age in years, mean | -0.089 | 0.031 | 1.000 | 9611 |
| Urban, No. | -0.056 | 0.198 | 1.000 | 10918 |
| *Traumatic Event* | -0.034 | 0.162 | 1.000 | 11320 |
| *Total Scores* | | | | |
| Depressed mood (QIDS-adjusted[†]) | 0.125 | 0.031 | 1.000 | 9759 |
| Anxiety (OASIS) | -0.024 | 0.024 | 1.000 | 10481 |
| Psychosis-like experiences (PQ16) | 0.055 | 0.023 | 1.000 | 9803 |
| Mania-like experiences (ASRM) | -0.042 | 0.026 | 1.000 | 11623 |
| Functioning (WSAS) | 0.076 | 0.014 | 1.000 | 9613 |
| Social support (SSSS) | 0.046 | 0.023 | 1.000 | 10949 |
| Sleep (BMC Sleep-Wake Scale) | -0.070 | 0.130 | 1.000 | 11300 |

[†] QIDS-adjusted is calculated as the total QIDS score with the value of question 12, which asks about suicidal ideation, subtracted.

### 4. Item-Level Undirected Probabilistic Graph Model Inference

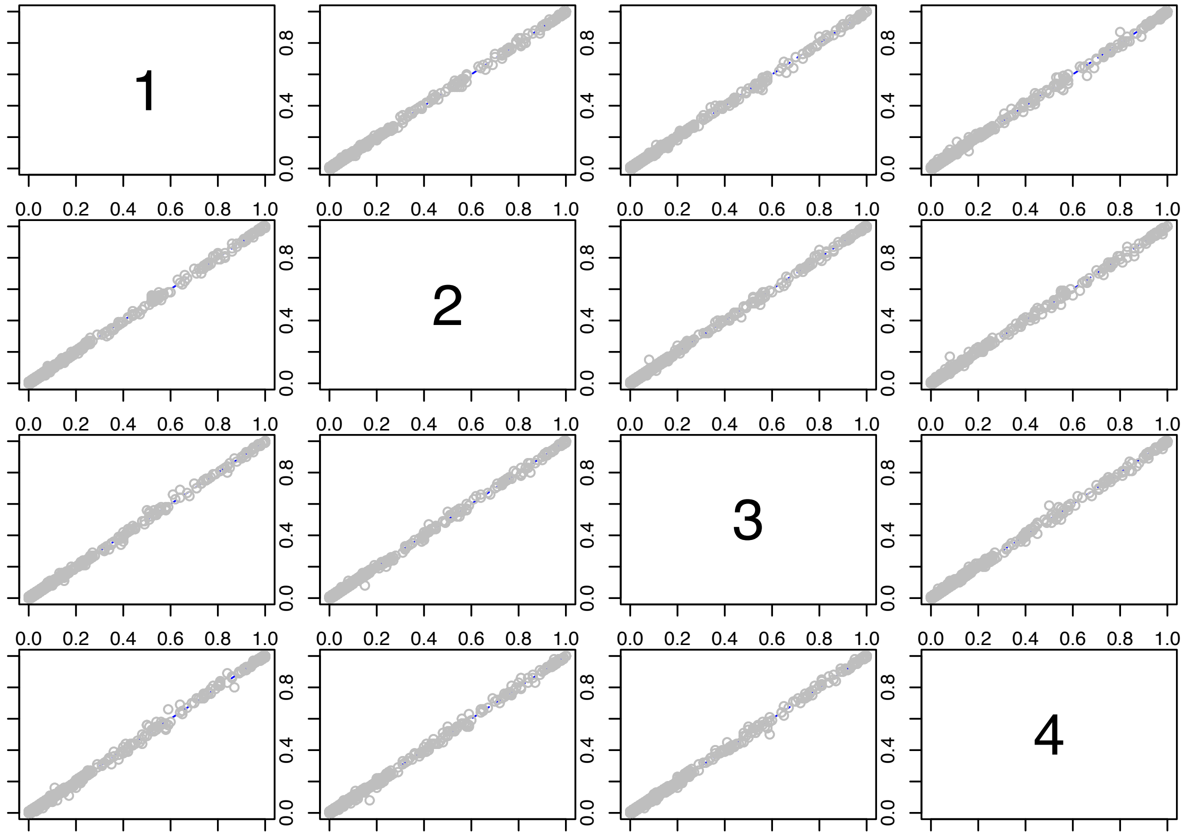

**Figure S1.** Concordance plot for the item-level undirected pairwise edge probabilities. We ensured that the difference in estimated edge probabilities between all chain pairs was below 0.1.

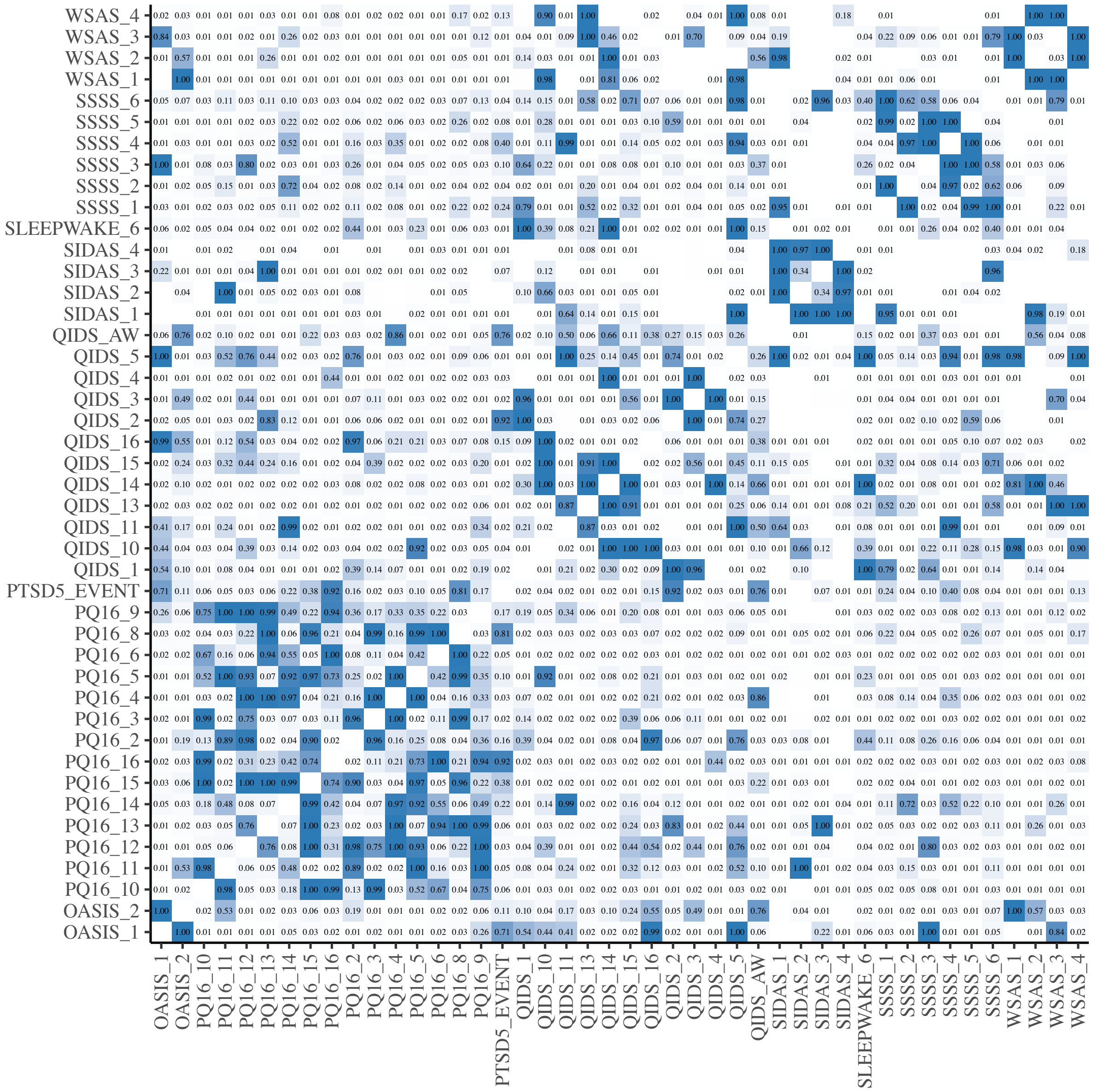

**Figure S2.** Pairwise edge probabilities for the undirected PGM.

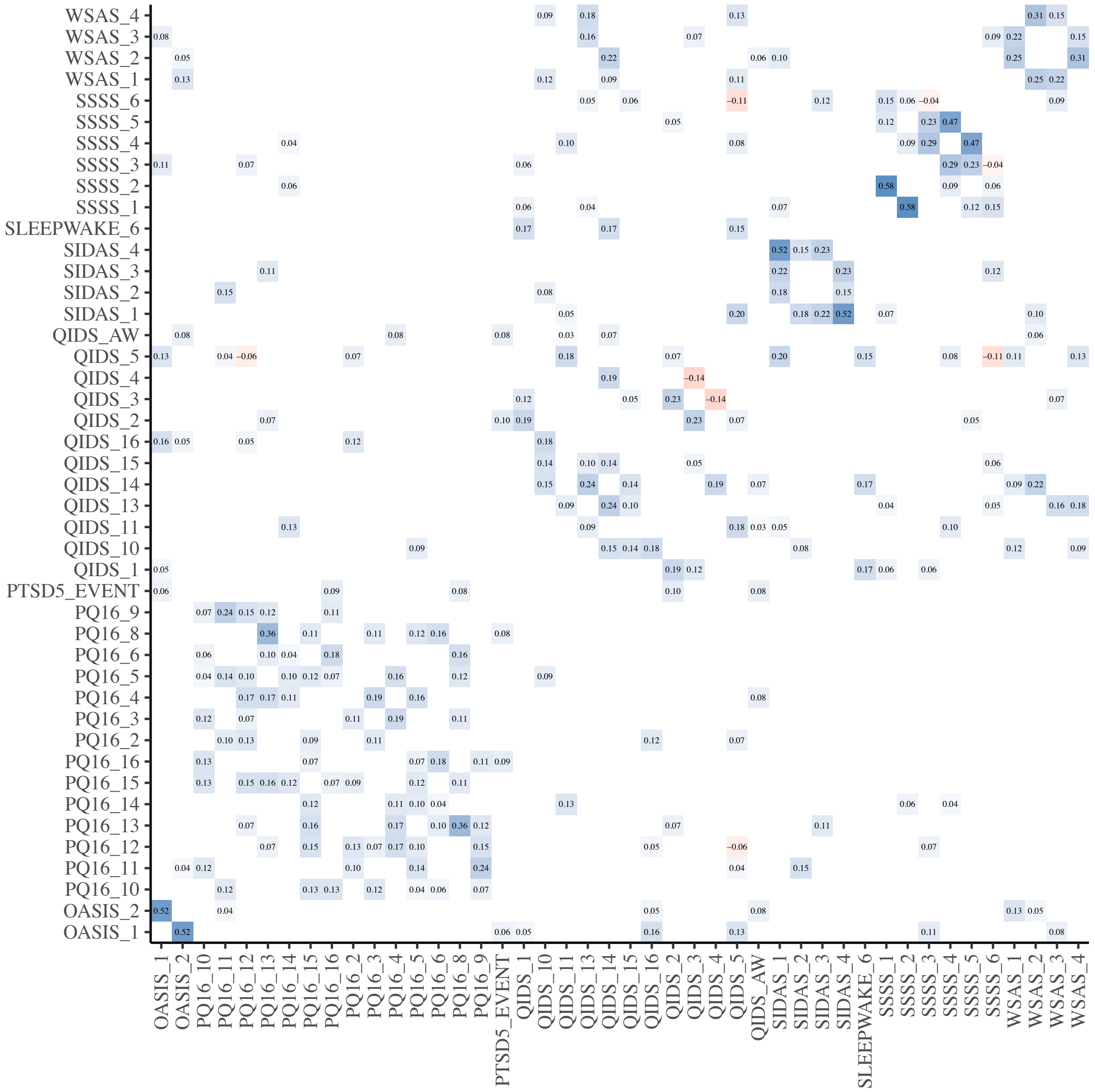

**Figure S3.** Partial correlations for edges with posterior probability greater than 0.5.

### 5. Factor-Level Bayesian Network Inference

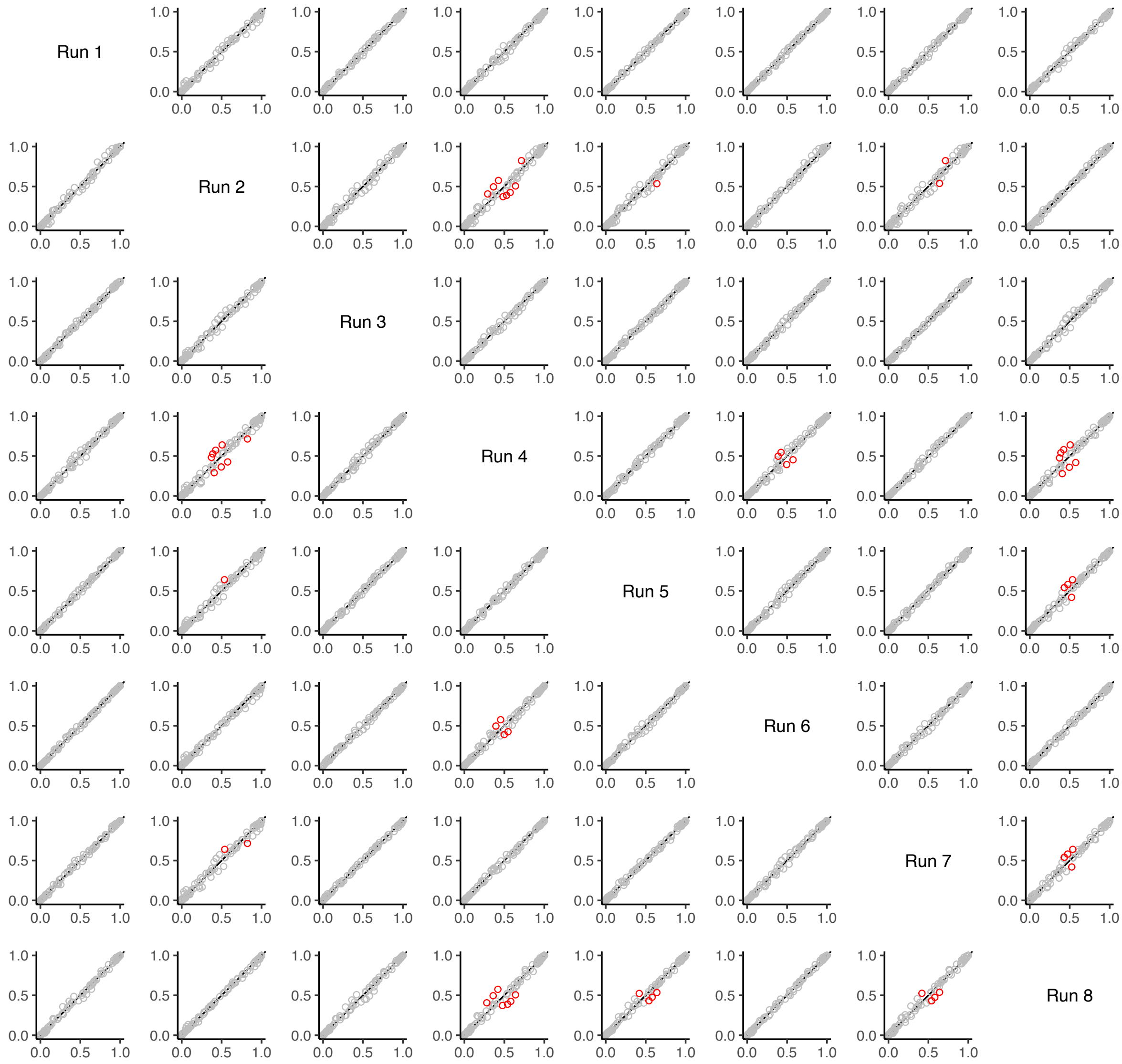

**Figure S4.** Concordance plot for the factor-level Bayesian network pairwise edge probabilities. We ensured that the difference in estimated edge probabilities between all chain pairs was below 0.2, but highlight differences between 0.1 and 0.2.

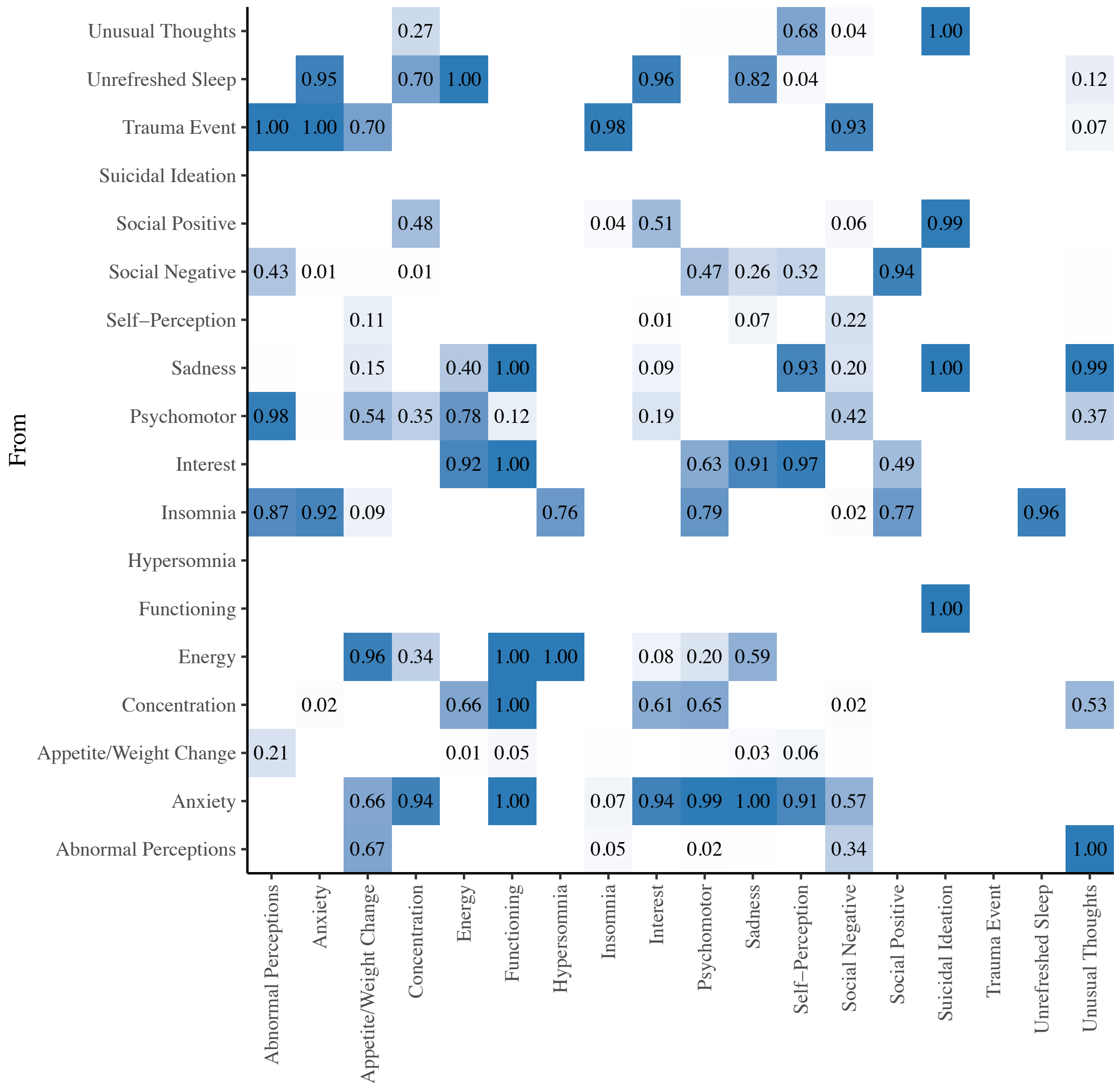

**Figure S5.** Pairwise edge probabilities where the edge points from row to column for the Bayesian network inference. We only show cases where the pairwise edge probabilities are greater than 0.

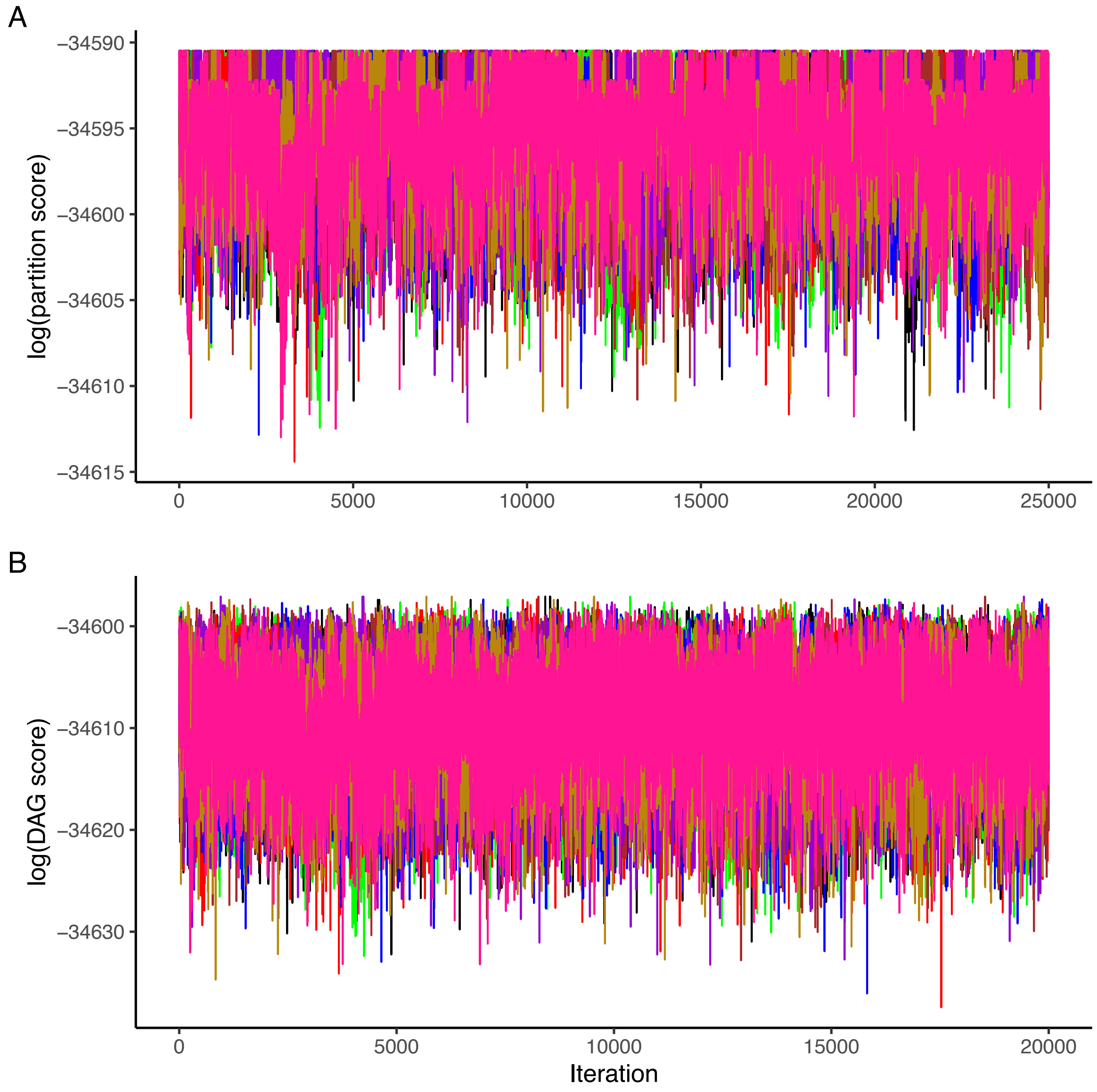

**Figure S6.** Trace plots for log scores in (A) partition and (B) DAG spaces.

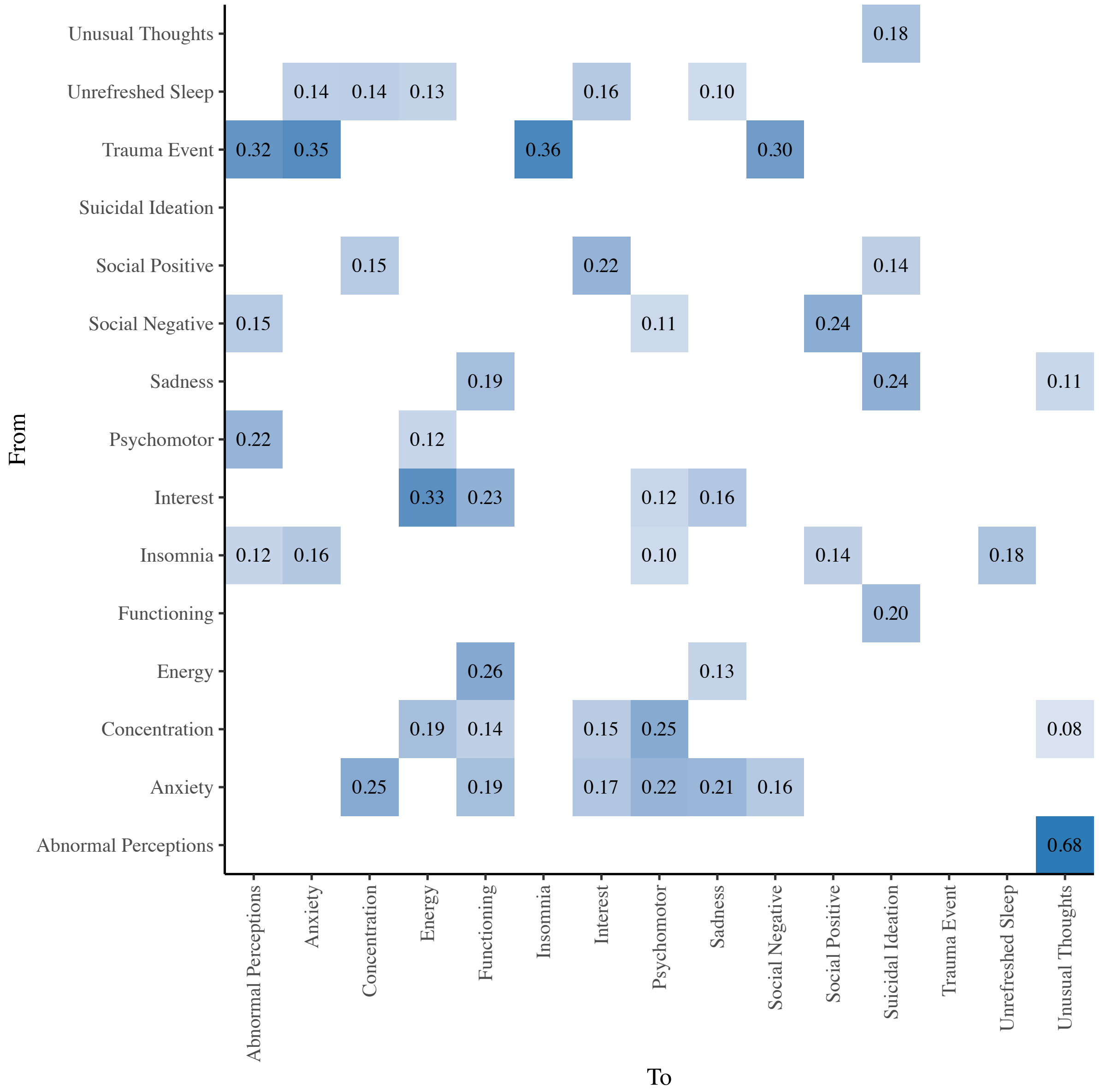

**Figure S7.** Linear regression coefficients estimated for the *maximum a posteriori* directed acyclic subgraph presented in Figure 2 of the manuscript. For ease of visualisation the fill colour property has been saturated at 0.4.

**Table S8.** Causal effect median, interquartile range, $\hat{R}$ convergence statistic, and effective sample sizes ($N_{\mathrm{eff}}$) for edges where zero is not within the IQR (Gelman et al., 2013; Vehtari et al., 2021).

| From | To | Median | Q25 | Q75 | $\hat{R}$ | $N_{\mathrm{eff}}$ |
|---|---|---|---|---|---|---|
| Abnormal Perceptions | Unusual Thoughts | 0.682 | 0.666 | 0.698 | 1.000 | 117250 |
| Abnormal Perceptions | Suicidal Ideation | 0.122 | 0.109 | 0.136 | 1.001 | 34382 |
| Anxiety | Abnormal Perceptions | 0.087 | 0.077 | 0.098 | 1.003 | 9133 |
| Anxiety | Unusual Thoughts | 0.111 | 0.100 | 0.122 | 1.003 | 8636 |
| Anxiety | Concentration | 0.242 | 0.219 | 0.264 | 1.001 | 13529 |
| Anxiety | Self-Perception | 0.217 | 0.191 | 0.24 | 1.000 | 64304 |
| Anxiety | Interest | 0.211 | 0.188 | 0.233 | 1.001 | 11572 |
| Anxiety | Energy | 0.152 | 0.136 | 0.167 | 1.002 | 9622 |
| Anxiety | Psychomotor | 0.33 | 0.308 | 0.351 | 1.003 | 8228 |
| Anxiety | Hypersomnia | 0.035 | 0.030 | 0.040 | 1.003 | 10117 |
| Anxiety | Sadness | 0.276 | 0.255 | 0.297 | 1.001 | 23128 |
| Anxiety | Appetite/Weight Change | 0.147 | 0.084 | 0.18 | 1.000 | 88672 |
| Anxiety | Suicidal Ideation | 0.164 | 0.153 | 0.176 | 1.001 | 17159 |
| Anxiety | Social Negative | 0.129 | 0.083 | 0.169 | 1.009 | 4881 |
| Anxiety | Social Positive | 0.052 | 0.038 | 0.071 | 1.003 | 6234 |
| Anxiety | Functioning | 0.354 | 0.334 | 0.374 | 1.001 | 18917 |
| Concentration | Suicidal Ideation | 0.077 | 0.034 | 0.093 | 1.002 | 5182 |
| Concentration | Functioning | 0.237 | 0.152 | 0.267 | 1.002 | 5833 |
| Energy | Unusual Thoughts | 0.017 | 0.009 | 0.025 | 1.002 | 5606 |
| Energy | Hypersomnia | 0.238 | 0.217 | 0.259 | 1.000 | 151980 |
| Energy | Appetite/Weight Change | 0.139 | 0.115 | 0.162 | 1.000 | 73569 |
| Energy | Suicidal Ideation | 0.082 | 0.058 | 0.100 | 1.003 | 5955 |
| Energy | Functioning | 0.281 | 0.257 | 0.308 | 1.003 | 7750 |
| Functioning | Suicidal Ideation | 0.203 | 0.182 | 0.223 | 1.000 | 159830 |
| Insomnia | Anxiety | 0.177 | 0.153 | 0.199 | 1.008 | 5713 |
| Insomnia | Abnormal Perceptions | 0.160 | 0.131 | 0.183 | 1.006 | 8629 |
| Insomnia | Unusual Thoughts | 0.127 | 0.106 | 0.144 | 1.007 | 7478 |
| Insomnia | Concentration | 0.084 | 0.069 | 0.098 | 1.007 | 4897 |
| Insomnia | Self-Perception | 0.058 | 0.049 | 0.067 | 1.008 | 5688 |
| Insomnia | Interest | 0.088 | 0.071 | 0.104 | 1.008 | 4081 |
| Insomnia | Energy | 0.086 | 0.074 | 0.096 | 1.009 | 4646 |
| Insomnia | Psychomotor | 0.173 | 0.129 | 0.199 | 1.007 | 7326 |
| Insomnia | Hypersomnia | -0.080 | -0.107 | -0.03 | 1.000 | 130165 |
| Insomnia | Sadness | 0.079 | 0.068 | 0.088 | 1.009 | 5067 |
| Insomnia | Appetite/Weight Change | 0.055 | 0.043 | 0.067 | 1.003 | 17131 |
| Insomnia | Suicidal Ideation | 0.078 | 0.067 | 0.087 | 1.009 | 4963 |
| Insomnia | Unrefreshed Sleep | 0.173 | 0.151 | 0.195 | 1.010 | 5300 |

| | | | | | | |
|---|---|---|---|---|---|---|
| Insomnia | Social Negative | 0.038 | 0.027 | 0.054 | 1.008 | 7510 |
| Insomnia | Social Positive | 0.134 | 0.082 | 0.16 | 1.003 | 9718 |
| Insomnia | Functioning | 0.102 | 0.090 | 0.113 | 1.009 | 4528 |
| Interest | Abnormal Perceptions | 0.026 | 0.007 | 0.038 | 1.003 | 6352 |
| Interest | Unusual Thoughts | 0.043 | 0.029 | 0.056 | 1.003 | 5991 |
| Interest | Self-Perception | 0.174 | 0.150 | 0.196 | 1.001 | 21473 |
| Interest | Energy | 0.357 | 0.329 | 0.384 | 1.003 | 4960 |
| Interest | Psychomotor | 0.113 | 0.036 | 0.157 | 1.003 | 5809 |
| Interest | Hypersomnia | 0.084 | 0.073 | 0.095 | 1.003 | 5869 |
| Interest | Sadness | 0.208 | 0.182 | 0.231 | 1.002 | 7515 |
| Interest | Appetite/Weight Change | 0.059 | 0.046 | 0.072 | 1.002 | 7773 |
| Interest | Suicidal Ideation | 0.141 | 0.124 | 0.162 | 1.003 | 7444 |
| Interest | Functioning | 0.360 | 0.336 | 0.382 | 1.002 | 6535 |
| Psychomotor | Abnormal Perceptions | 0.238 | 0.216 | 0.259 | 1.003 | 7693 |
| Psychomotor | Unusual Thoughts | 0.183 | 0.159 | 0.227 | 1.002 | 10879 |
| Psychomotor | Self-Perception | 0.023 | 0.010 | 0.035 | 1.005 | 7095 |
| Psychomotor | Energy | 0.139 | 0.081 | 0.177 | 1.004 | 5323 |
| Psychomotor | Hypersomnia | 0.032 | 0.018 | 0.042 | 1.004 | 5677 |
| Psychomotor | Appetite/Weight Change | 0.099 | 0.051 | 0.157 | 1.000 | 63877 |
| Psychomotor | Suicidal Ideation | 0.053 | 0.039 | 0.067 | 1.005 | 6044 |
| Psychomotor | Functioning | 0.068 | 0.028 | 0.093 | 1.004 | 7075 |
| Sadness | Unusual Thoughts | 0.114 | 0.099 | 0.130 | 1.001 | 17509 |
| Sadness | Self-Perception | 0.215 | 0.189 | 0.239 | 1.001 | 17375 |
| Sadness | Suicidal Ideation | 0.304 | 0.284 | 0.324 | 1.000 | 82545 |
| Sadness | Functioning | 0.219 | 0.198 | 0.241 | 1.000 | 19690 |
| Social Negative | Suicidal Ideation | 0.048 | 0.032 | 0.071 | 1.008 | 4755 |
| Social Negative | Social Positive | 0.235 | 0.211 | 0.257 | 1.001 | 17049 |
| Social Positive | Suicidal Ideation | 0.161 | 0.136 | 0.186 | 1.003 | 9418 |
| Trauma Event | Anxiety | 0.394 | 0.351 | 0.436 | 1.000 | 101751 |
| Trauma Event | Abnormal Perceptions | 0.502 | 0.463 | 0.542 | 1.000 | 82411 |
| Trauma Event | Unusual Thoughts | 0.372 | 0.342 | 0.403 | 1.000 | 67168 |
| Trauma Event | Insomnia | 0.358 | 0.315 | 0.400 | 1.002 | 26355 |
| Trauma Event | Concentration | 0.124 | 0.106 | 0.143 | 1.001 | 10565 |
| Trauma Event | Self-Perception | 0.127 | 0.109 | 0.145 | 1.001 | 26939 |
| Trauma Event | Interest | 0.110 | 0.092 | 0.127 | 1.005 | 6098 |
| Trauma Event | Energy | 0.087 | 0.075 | 0.099 | 1.003 | 7666 |
| Trauma Event | Psychomotor | 0.188 | 0.164 | 0.212 | 1.002 | 9958 |
| Trauma Event | Sadness | 0.128 | 0.111 | 0.147 | 1.002 | 15937 |
| Trauma Event | Appetite/Weight Change | 0.273 | 0.165 | 0.33 | 1.000 | 116029 |
| Trauma Event | Suicidal Ideation | 0.146 | 0.133 | 0.161 | 1.002 | 17762 |
| Trauma Event | Unrefreshed Sleep | 0.061 | 0.05 | 0.072 | 1.007 | 7333 |
| Trauma Event | Social Negative | 0.358 | 0.308 | 0.404 | 1.002 | 13968 |

| | | | | | | |
|---|---|---|---|---|---|---|
| Trauma Event | Social Positive | 0.129 | 0.105 | 0.15 | 1.002 | 11723 |
| Trauma Event | Functioning | 0.162 | 0.143 | 0.181 | 1.002 | 17583 |
| Unrefreshed Sleep | Anxiety | 0.135 | 0.113 | 0.157 | 1.001 | 28411 |
| Unrefreshed Sleep | Abnormal Perceptions | 0.023 | 0.018 | 0.028 | 1.004 | 7374 |
| Unrefreshed Sleep | Unusual Thoughts | 0.048 | 0.039 | 0.058 | 1.000 | 44089 |
| Unrefreshed Sleep | Concentration | 0.153 | 0.103 | 0.182 | 1.001 | 8043 |
| Unrefreshed Sleep | Self-Perception | 0.083 | 0.071 | 0.094 | 1.000 | 61794 |
| Unrefreshed Sleep | Interest | 0.206 | 0.183 | 0.227 | 1.000 | 28785 |
| Unrefreshed Sleep | Energy | 0.242 | 0.222 | 0.263 | 1.000 | 84811 |
| Unrefreshed Sleep | Psychomotor | 0.090 | 0.074 | 0.105 | 1.003 | 9592 |
| Unrefreshed Sleep | Hypersomnia | 0.057 | 0.05 | 0.064 | 1.000 | 111956 |
| Unrefreshed Sleep | Sadness | 0.184 | 0.15 | 0.208 | 1.000 | 58130 |
| Unrefreshed Sleep | Appetite/Weight Change | 0.054 | 0.045 | 0.062 | 1.000 | 45072 |
| Unrefreshed Sleep | Suicidal Ideation | 0.094 | 0.081 | 0.105 | 1.000 | 27941 |
| Unrefreshed Sleep | Social Negative | 0.024 | 0.018 | 0.031 | 1.001 | 12400 |
| Unrefreshed Sleep | Social Positive | 0.010 | 0.005 | 0.055 | 1.010 | 3219 |
| Unrefreshed Sleep | Functioning | 0.187 | 0.171 | 0.203 | 1.000 | 33971 |
| Unusual Thoughts | Suicidal Ideation | 0.177 | 0.158 | 0.196 | 1.000 | 59206 |

### 6. References

Altman, E. G., Hedeker, D., Peterson, J. L., & Davis, J. M. (1997). The altman self-rating Mania scale. *Biological Psychiatry*, *42*(10), 948–955. https://doi.org/10.1016/S0006-3223(96)00548-3

Andrews, G., & Slade, T. (2001). Interpreting scores on the Kessler Psychological Distress Scale (K10). *Australian and New Zealand Journal of Public Health*, *25*(6), 494–497. https://doi.org/10.1111/j.1467-842X.2001.tb00310.x

Babor, T. F., Higgins-Biddle, J. C., Saunders, J. B., & Monteiro, M. G. (2001). AUDIT: the Alcohol Use Disorders Identification Test: guidelines for use in primary health care. In *Guidelines for Use in Primary Care (second edition)*.

Brown, E. S., Murray, M., Carmody, T. J., Kennard, B. D., Hughes, C. W., Khan, D. A., & Rush, A. J. (2008). The Quick Inventory of Depressive Symptomatology-Self-report: A psychometric evaluation in patients with asthma and major depressive disorder. *Annals of Allergy, Asthma and Immunology*, *100*(5), 433–438. https://doi.org/10.1016/S1081-1206(10)60467-X

Bush, K., Kivlahan, D. R., McDonnell, M. B., Fihn, S. D., Bradley, K. A., & (ACQUIP), for the A. C. Q. I. P. (1998). The AUDIT Alcohol Consumption Questions (AUDIT-C): An Effective Brief Screening Test for Problem Drinking. *Arch Intern Med.*, *158*(16), 1789–1795. https://doi.org/10.1001/archinte.158.16.1789

Buysse, D. J., Reynolds, C. F., Monk, T. H., Berman, S. R., & Kupfer, D. J. (1989). The Pittsburgh Sleep Quality Index: a new instrument for psychiatric practice and research. Psychiatry Res. 1989;28:193–213. *Psychiatry Research*, *28*(2), 193–213. https://doi.org/10.1016/0165-1781(89)90047-4

Campbell-Sills, L., Norman, S. B., Craske, M. G., Sullivan, G., Lang, A. J., Chavira, D. A., Bystritsky, A., Sherbourne, C., Roy-Byrne, P., & Stein, M. B. (2009). Validation of a brief measure of anxiety-related severity and impairment: The Overall Anxiety Severity and Impairment Scale (OASIS). *Journal of Affective Disorders*, *112*(1–3), 92–101. https://doi.org/10.1016/j.jad.2008.03.014

Capon, W., Hickie, I. B., McKenna, S., Varidel, M., Richards, M., LaMonica, H. M., Rock, D., Scott, E. M., & Iorfino, F. (2023). Characterising variability in youth mental health service populations: A detailed and scalable approach using digital technology. *Australasian Psychiatry*, *31*(3), 295–301. https://doi.org/10.1177/10398562231167681

Craig, C. L., Marshall, A. L., Sjöström, M., Bauman, A. E., Booth, M. L., Ainsworth, B. E., Pratt, M., Ekelund, U., Yngve, A., Sallis, J. F., & Oja, P. (2003). International physical activity questionnaire: 12-Country reliability and validity. *Medicine and Science in Sports and Exercise*, *35*(8), 1381–1395. https://doi.org/10.1249/01.MSS.0000078924.61453.FB

Department of Health and Aged Care. (2021). *Body mass index (BMI) and waist measurement*. https://www.health.gov.au/topics/overweight-and-obesity/bmi-and-waist

Gelman, A., Carlin, J. B., Stern, H. S., Dunson, D. B., Vehtari, A., & Rubin, D. B. (2013). *Bayesian Data Analysis* (3rd Ed.). Chapman and Hall/CRC. https://doi.org/10.1201/9780429258411

Goldman, H. H., Skodol, A. E., & Lave, T. R. (1992). Revising axis V for DSM-IV: A review of measures of social functioning. *American Journal of Psychiatry*, *149*(9), 1148–1156. https://doi.org/10.1176/ajp.149.9.1148

Goodrich, B., Gabry, J., Ali, A., & Brilleman, S. (2024). *rstanarm: Bayesian applied regression modeling via Stan* (R package version 2.32.1). https://mc-stan.org/rstanarm

Hay, P. J., Mond, J., Buttner, P., & Darby, A. (2008). Eating disorder behaviors are increasing: Findings from two sequential community surveys in South Australia. *PLoS ONE*, *3*(2), 1–5. https://doi.org/10.1371/journal.pone.0001541

Howie, C., Hanna, D., Shannon, C., Davidson, G., & Mulholland, C. (2022). The Structure of the Prodromal Questionnaire-16 (PQ-16): Exploratory and confirmatory factor analyses in a general non-help-seeking population sample. *Early Intervention in Psychiatry*, *16*(3), 239–246. https://doi.org/10.1111/eip.13147

Humeniuk, R., Ali, R., Babor, T. F., Farrell, M., Formigoni, M. L., Jittiwutikarn, J., De Lacerda, R. B., Ling, W., Marsden, J., Monteiro, M., Nhiwatiwa, S., Pal, H., Poznyak, V., & Simon, S. (2008). Validation of the alcohol, smoking and substance involvement screening test (ASSIST). *Addiction*, *103*(6), 1039–1047. https://doi.org/10.1111/j.1360-0443.2007.02114.x

Ising, H. K., Veling, W., Loewy, R. L., Rietveld, M. W., Rietdijk, J., Dragt, S., Klaassen, R. M. C., Nieman, D. H., Wunderink, L., Linszen, D. H., & Van Der Gaag, M. (2012). The validity of the 16-item version of the prodromal questionnaire (PQ-16) to screen for ultra high risk of developing psychosis in the general help-seeking population. *Schizophrenia Bulletin*, *38*(6), 1288–1296. https://doi.org/10.1093/schbul/sbs068

Kessler, R. C., Andrews, G., Colpe, L. J., Hiripi, E., Mroczek, D. K., Normand, S. L. T., Walters, E. E., & Zaslavsky, A. M. (2002). Short screening scales to monitor population prevalences and trends in non-specific psychological distress. *Psychological Medicine*, *32*(6), 959–976. https://doi.org/10.1017/S0033291702006074

Mundt, J. C., Marks, I. M., Shear, M. K., & Greist, J. H. (2002). The Work and Social Adjustment Scale: A simple measure of impairment in functioning. *British Journal of Psychiatry*, *180*(MAY), 461–464. https://doi.org/10.1192/bjp.180.5.461

Norman, S. B., Cissell, S. H., Means-Christensen, A. J., & Stein, M. B. (2006). Development and Validation of an Overall Anxiety Severity and Impairment Scale (OASIS). *Depression and Anxiety*, *23*(4), 245–249. https://doi.org/10.1002/da.20182

OECD. (2023). *Youth not in employment, education or training (NEET)*. https://doi.org/https://doi.org/10.1787/72d1033a-en

Posner, K., Brown, G. K., & Stanley, B. (2011). The Columbia–Suicide Severity Rating Scale: Initial Validity and Internal Consistency Findings From Three Multisite Studies With Adolescents and Adults. *American Journal of Psychiatry*, *168*(12), 1267–1277. https://doi.org/10.1176/appi.ajp.2011.10111704

Prins, A., Bovin, M. J., Smolenski, D. J., Marx, B. P., Kimerling, R., Jenkins-Guarnieri, M. A., Kaloupek, D. G., Schnurr, P. P., Kaiser, A. P., Leyva, Y. E., & Tiet, Q. Q. (2016). The Primary Care PTSD Screen for DSM-5 (PC-PTSD-5): Development and Evaluation Within a Veteran Primary Care Sample. *Journal of General Internal Medicine*, *31*(10), 1206–1211. https://doi.org/10.1007/s11606-016-3703-5

Roenneberg, T., Wirz-Justice, A., & Merrow, M. (2003). Life between clocks: Daily temporal patterns of human chronotypes. *Journal of Biological Rhythms*, *18*(1), 80–90. https://doi.org/10.1177/0748730402239679

Rush, A. J., Trivedi, M. H., Ibrahim, H. M., Carmody, T. J., Arnow, B., Klein, D. N., Markowitz, J. C., Ninan, P. T., Kornstein, S., Manber, R., Thase, M. E., Kocsis, J. H., & Keller, M. B. (2003). The 16-item Quick Inventory of Depressive Symptomatology (QIDS), clinician rating (QIDS-C), and self-report (QIDS-SR): A psychometric evaluation in patients with chronic major depression. *Biological Psychiatry*, *54*(5), 573–583. https://doi.org/10.1016/S0006-3223(02)01866-8

Schuster, T. L., Kessler, R. C., & Aseltine, R. H. (1990). Supportive interactions, negative interactions, and depressed mood. *American Journal of Community Psychology*, *18*(3), 423–438. https://doi.org/10.1007/BF00938116

Van Spijker, B. A. J., Batterham, P. J., Calear, A. L., Farrer, L., Christensen, H., Reynolds, J., & Kerkhof, A. J. F. M. (2014). The Suicidal Ideation Attributes Scale (SIDAS): Community-based validation study of a new scale for the measurement of suicidal ideation. *Suicide and Life-Threatening Behavior*, *44*(4), 408–419. https://doi.org/10.1111/sltb.12084

Vehtari, A., Gelman, A., Simpson, D., Carpenter, B., & Burkner, P. C. (2021). Rank-Normalization, Folding, and Localization: An Improved R-hat for Assessing Convergence of MCMC (with Discussion). *Bayesian Analysis*, *16*(2), 667–718. https://doi.org/10.1214/20-BA1221
